# Single-nucleus transcriptomics reveals disease- and pathology-specific signatures in α-synucleinopathies

**DOI:** 10.1101/2023.10.10.23296642

**Authors:** Gonzalo S Nido, Martina Castelli, Sepideh Mostafavi, Anna Rubiolo, Omnia Shadad, Guido Alves, Ole-Bjørn Tysnes, Christian Dölle, Charalampos Tzoulis

## Abstract

α-synucleinopathies are severe neurodegenerative disorders characterized by intracellular aggregation of α-synuclein, yet their molecular pathogenesis remains unknow. Here, we explore cell-specific changes in gene expression across different α-synucleinopathies. We perform single-nucleus RNA sequencing (snRNA-seq) on nearly 300,000 nuclei from the prefrontal cortex of individuals with idiopathic Parkinson’s disease (iPD), Parkinson’s disease caused by *LRRK2* mutations (LRRK2-PD), multiple system atrophy (MSA) and healthy controls. iPD and LRRK2-PD exhibit a largely overlapping cell type-specific signature, which is distinct from that of MSA, and includes an overall decrease of the transcriptional output in neurons. Notably, most of the differential expression signal in iPD and LRRK2-PD is concentrated in a specific deep cortical neuronal subtype expressing adrenoceptor alpha 2A. While most differentially expressed genes are highly cell type- and disease-specific, *PDE10A* is found consistently downregulated in most cortical neurons, and across all three diseases. Finally, exploiting the variable presence and/or severity of α-synuclein pathology in LRRK2-PD and iPD, we identify cell type-specific signatures associated with α-synuclein pathology, including a neuronal upregulation of the *SNCA* gene itself, encoding α-synuclein. Our findings provide novel insights into the cell-specific transcriptional landscape of the α-synucleinopathy spectrum.

## Introduction

α-synucleinopathies are relentlessly progressive, debilitating, and incurable neurodegenerative disorders comprising Parkinson’s disease (PD), dementia with Lewy bodies (DLB) and multiple system atrophy (MSA)^1,2^. These disorders share the common feature of intracellular α-synuclein-positive inclusions, which are generally believed to contribute to the neurodegenerative process^3^. In PD and DLB, α-synuclein inclusions occur mostly in neurons, assuming the form of Lewy bodies and neurites, collectively termed Lewy pathology, whereas in MSA they predominantly affect oligodendrocytes in the form of oligodendroglial cytoplasmic inclusions^4,5^. While α-synuclein pathology is a *sine qua non* feature of MSA, DLB, and idiopathic PD (iPD), it is not always present in monogenic forms of PD, such as in patients with *LRRK2* mutations, about half of whom exhibit no Lewy pathology^6,7^. Currently, there are no disease-modifying therapies for α-synucleinopathies and clinical trials have produced disappointing results. This is mainly attributed to the lack of mechanistic insight into the molecular processes underlying neuronal dysfunction and -death in these diseases.

Whole-genome transcriptomic studies in brain tissue from individuals with α-synucleinopathies have the potential to provide valuable new insights in disease-associated changes in gene expression. However, prior studies have shown surprisingly low replicability at the gene level and only partial concordance at the pathway level, which appears to be largely driven by differences in cell composition (i.e., neuronal loss and gliosis), rather than disease-related changes within cells^8–10^. Single-nucleus RNA sequencing (snRNA-seq) has emerged as a powerful approach to mitigate this limitation allowing the identification of cell type-specific changes in gene expression. The first snRNA-seq studies on α-synucleinopathies have focused on the dopaminergic *substantia nigra pars compacta* (SNc) in PD, providing unprecedented insights into the transcriptional state of individual cells^11,12^. One important drawback of studying SNc tissue is that it is typically severely degenerated in the terminal stage of the disease, exhibiting loss of approximately 80% of the dopaminergic neuronal population on average^13^. This greatly complicates the comparison between cases and controls, as surviving dopaminergic neurons are difficult to isolate in sufficient numbers and are likely to be resilient cells, introducing survival bias. For this reason, we believe that regions with relatively milder and later involvement, such as the neocortex, may be more informative in terms of disease processes involved in early cell-specific dysfunction and dysregulation.

Here, we employed snRNA-seq to identify cell type proportions and cell type-specific gene expression signatures in the prefrontal cortex of individuals with iPD, monogenic PD caused by *LRRK2* mutations (LRRK2-PD), and MSA, compared to demographically matched controls. We find that the cell type-specific signature of iPD and LRRK-PD are qualitatively similar and distinct from that of MSA, and identify a large number of disease- and cell type-specific differentially expressed genes. Furthermore, by comparing LRRK2-PD samples with and without α-synuclein pathology, we discern cell type-specific signatures of α-synuclein pathology.

## Results

### snRNA-seq profiling of the prefrontal cortex in α-synucleinopathies

We purified and isolated single nuclei from the prefrontal cortex (PFC) of individuals with iPD (n = 20), LRRK2-PD (n = 7), and parkinsonian type MSA (n = 6), as well as demographically matched neurologically healthy controls (n = 13; Figure 1A, Supp. Table 1). snRNA-sequencing data was obtained from a total of 299,582 high quality nuclei (see Methods). Integration of the complete dataset evidenced clusters for the main cortical cell types, including excitatory neurons, inhibitory neurons, oligodendrocytes, astrocytes, oligodendrocyte precursor cells (OPCs), endothelial cells and microglia, which could be identified by their respective marker expression levels (Figure 1C). High resolution clustering of the snRNA-seq dataset revealed a total of 28 subclusters: 12 excitatory neuronal, 8 inhibitory neuronal, 3 oligodendrocyte, 2 astrocyte, 1 microglial, 1 OPC, and 1 endothelial cluster (Figure 1D). Neuronal subclusters were further characterized by known markers of cortical neurons (Figure 1E-F). Conventional bulk RNA sequencing (RNA-seq) data was obtained from immediately adjacent cortical samples from the same individuals, except for 2 control samples that failed quality control (Supp. Table 1).

**Figure 1.**
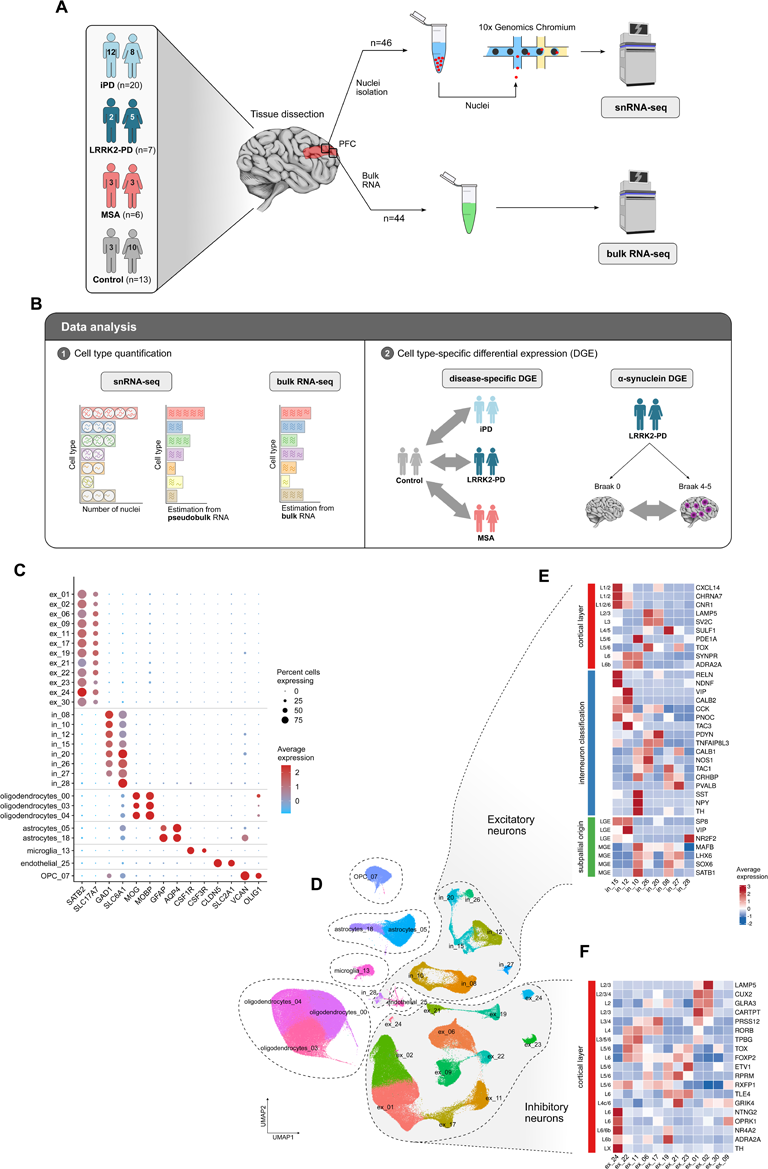
Single-nucleus transcriptomics in α-syncleinopathies. **(A)** Experimental workflow for tissue processing. Two adjacent tissue samples from fresh frozen PFC (Brodmann area 9) were obtained from 46 individuals. Nuclei suspensions were processed using the 10x Genomics Chromium platform and subsequently underwent deep RNA-sequencing (snRNA-seq). RNA-sequencing was also conducted in bulk tissue samples (bulk RNA-seq). **(B)** Both snRNA-seq and bulk RNA-seq datasets were assessed for cell type composition and differential gene expression. Cell type composition was estimated in the snRNA-seq dataset by two alternative approaches, based on the nuclei counts, or marker gene profiles^14^ (MGPs) on the pseudobulk data (see Methods). MGPs were also used to estimate cell types in the bulk RNA-seq samples. Differential gene expression was calculated for each disease group (i.e., control *vs* iPD, control *vs* LRRK2-PD and control *vs* MSA) and for Lewy pathology Braak stage 0 *vs* Braak stage 4-5 within the subset of LRRK2-PD samples. Grey arrows indicate the contrasts. **(C)** Dot plot of average gene expression in subclusters of cell type markers for the main cortical cell types. **(D)** UMAP embeddings for the snRNA-seq dataset. Colors correspond to different clusters identified by Seurat at a resolution of 0.5. Dashed contours are drawn to aid visualization of the clusters corresponding to the main cortical cell types (excitatory neurons, inhibitory neurons, oligodendrocytes, astrocytes, microglia, oligodendrocyte precursor cells (OPCs) and endothelial cells. **(E)** Heatmap of average expression of markers for excitatory neurons. Left annotation labels indicate cortical layer (red), interneuron classification (blue) and subpallial origin (green) as proposed by Lake *et. al*^51^. **(F)** Heatmap of average expression of markers for inhibitory cortical neurons. Labels on the left-hand side indicate the markers’ cortical layer location^51^.

**Table 1.**
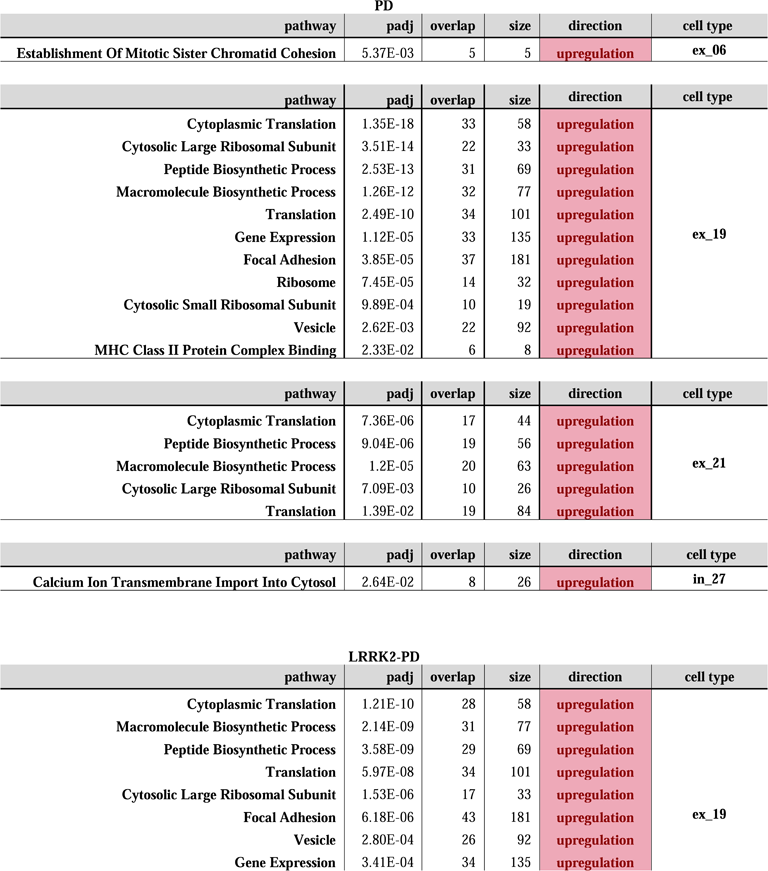
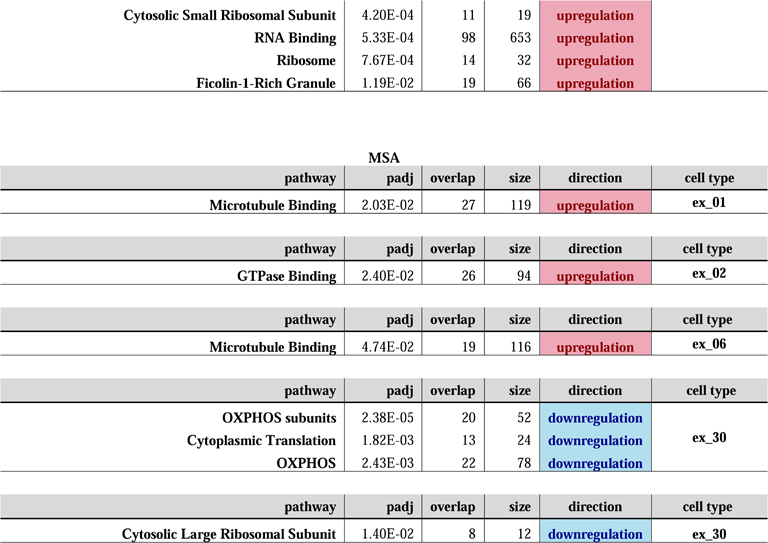
Functional overrepresentation analysis of differential expression in α-synucleinopathies. Significantly enriched GO terms for each of the disease groups. Padj: Benjamini-Hochberg adjusted p; overlap: DEGs overlapping the gene list; size: size of the gene list; direction: direction of change of enriched DEGs.

| PD |  |  |  |  |  |
| --- | --- | --- | --- | --- | --- |
| pathway | padj | overlap | size | direction | cell type |
| Establishment Of Mitotic Sister Chromatid Cohesion | 5.37E-03 | 5 | 5 | upregulation | ex_06 |
| pathway | padj | overlap | size | direction | cell type |
| Cytoplasmic Translation | 1.35E-18 | 33 | 58 | upregulation | ex_19 |
| Cytosolic Large Ribosomal Subunit | 3.51E-14 | 22 | 33 | upregulation |  |
| Peptide Biosynthetic Process | 2.53E-13 | 31 | 69 | upregulation |  |
| Macromolecule Biosynthetic Process | 1.26E-12 | 32 | 77 | upregulation |  |
| Translation | 2.49E-10 | 34 | 101 | upregulation |  |
| Gene Expression | 1.12E-05 | 33 | 135 | upregulation |  |
| Focal Adhesion | 3.85E-05 | 37 | 181 | upregulation |  |
| Ribosome | 7.45E-05 | 14 | 32 | upregulation |  |
| Cytosolic Small Ribosomal Subunit | 9.89E-04 | 10 | 19 | upregulation |  |
| Vesicle | 2.62E-03 | 22 | 92 | upregulation |  |
| MHC Class II Protein Complex Binding | 2.33E-02 | 6 | 8 | upregulation |  |
| pathway | padj | overlap | size | direction | cell type |
| Cytoplasmic Translation | 7.36E-06 | 17 | 44 | upregulation | ex_21 |
| Peptide Biosynthetic Process | 9.04E-06 | 19 | 56 | upregulation |  |
| Macromolecule Biosynthetic Process | 1.2E-05 | 20 | 63 | upregulation |  |
| Cytosolic Large Ribosomal Subunit | 7.09E-03 | 10 | 26 | upregulation |  |
| Translation | 1.39E-02 | 19 | 84 | upregulation |  |
| pathway | padj | overlap | size | direction | cell type |
| Calcium Ion Transmembrane Import Into Cytosol | 2.64E-02 | 8 | 26 | upregulation | in_27 |
| LRRK2-PD |  |  |  |  |  |
| pathway | padj | overlap | size | direction | cell type |
| Cytoplasmic Translation | 1.21E-10 | 28 | 58 | upregulation | ex_19 |
| Macromolecule Biosynthetic Process | 2.14E-09 | 31 | 77 | upregulation |  |
| Peptide Biosynthetic Process | 3.58E-09 | 29 | 69 | upregulation |  |
| Translation | 5.97E-08 | 34 | 101 | upregulation |  |
| Cytosolic Large Ribosomal Subunit | 1.53E-06 | 17 | 33 | upregulation |  |
| Focal Adhesion | 6.18E-06 | 43 | 181 | upregulation |  |
| Vesicle | 2.80E-04 | 26 | 92 | upregulation |  |
| Gene Expression | 3.41E-04 | 34 | 135 | upregulation |  |

|  |  |  |  |  |
| --- | --- | --- | --- | --- |
| Cytosolic Small Ribosomal Subunit | 4.20E-04 | 11 | 19 | upregulation |
| RNA Binding | 5.33E-04 | 98 | 653 | upregulation |
| Ribosome | 7.67E-04 | 14 | 32 | upregulation |
| Ficolin-1-Rich Granule | 1.19E-02 | 19 | 66 | upregulation |

| MSA |  |  |  |  |  |
| --- | --- | --- | --- | --- | --- |
| pathway | padj | overlap | size | direction | cell type |
| Microtubule Binding | 2.03E-02 | 27 | 119 | upregulation | ex_01 |
| pathway | padj | overlap | size | direction | cell type |
| GTPase Binding | 2.40E-02 | 26 | 94 | upregulation | ex_02 |
| pathway | padj | overlap | size | direction | cell type |
| Microtubule Binding | 4.74E-02 | 19 | 116 | upregulation | ex_06 |
| pathway | padj | overlap | size | direction | cell type |
| OXPHOS subunits | 2.38E-05 | 20 | 52 | downregulation | ex_30 |
| Cytoplasmic Translation | 1.82E-03 | 13 | 24 | downregulation |  |
| OXPHOS | 2.43E-03 | 22 | 78 | downregulation |  |
| pathway | padj | overlap | size | direction | cell type |
| Cytosolic Large Ribosomal Subunit | 1.40E-02 | 8 | 12 | downregulation | ex_30 |

### Cortical neurons show lower transcriptional output in α-synucleinopathies

We assessed cell type composition in the PFC of cases and controls using multiple methods in both bulk RNA-seq and snRNA-seq data from the same individuals. In bulk RNA-seq data, cell composition was estimated using marker gene profiles (MGP) as previously described^14^. As expected, we observed trends for a relative decrease in the neuronal fraction and a concomitant increase in the glial cell populations in all three diseases, with the single exception of decreased oligodendrocyte fraction in MSA (Figure 2A). To calculate cell type estimates in the snRNA-seq, we aggregated raw reads from the filtered barcodes in each individual sample (i.e., pseudobulk data) and then calculated MGPs. Cellularity estimates from pseudobulk did not recapitulate the differences observed in bulk tissue, revealing no differences in the relative abundances of cell types (Figure 2B). Furthermore, the proportions of cell type counts based on the number of captured nuclei showed no major differences across the 28 identified cell types. In fact, most neuronal clusters exhibited a mild trend for increase in iPD, LRRK2-PD and MSA compared to controls, although this only reached significance for very few clusters and only at a nominal level (Figure 2C).

**Figure 2.**
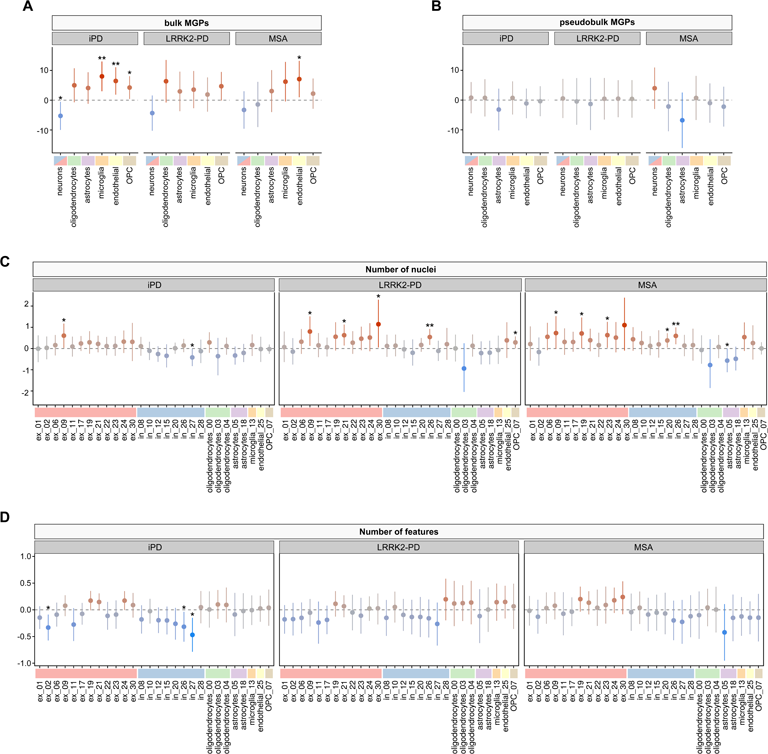
Cell type composition. Changes in relative cell type abundance are displayed as average fold-change in the disease compared to control samples. **(A)** MGP-based estimates calculated in bulk RNA-seq samples for the major cortical cell types showed a decrease in the relative abundance of neuronal transcripts and an increase in glial transcripts consistent with previous results. **(B)** MGP-based estimates calculated in pseudobulk tissue from the snRNA-seq samples (i.e., from a highly enriched nuclear fraction) of the major cortical cell types. Here, the cell type estimates did not exhibit significant deviations between cases and controls. **(C)** Changes in the proportions of captured nuclei in the snRNA-seq dataset for all cell type clusters. Similar to the pseudobulk results, the proportion of nuclei for the different cell types did not seem to follow the established notion of neuronal loss accompanied by gliosis, as observed in the bulk tissue transcriptome. On the contrary, nuclei of excitatory neurons appear overrepresented in the diseases. **(D)** Log fold-changes in the number of features (genes) detected per barcode show a mild but consistent decrease in the transcriptional output of most neuronal clusters compared to the glial populations in both iPD and LRRK2-PD. Coefficients were estimated as follows: for **A,B** (fold changes) with a linear regression; for **C** (log fold-changes) using a negative binomial regression with (log) total number of nuclei as offset; for **D** (log fold-changes) using a linear mixed model with sample as random effect. For all models, sex, age, and PMI were accounted for in the fit. Vertical bars: 95% confidence interval; *p < 0.05; **p < 0.01.

While there is evidence of bias towards certain cell type populations during nuclei isolation^15^, it is unlikely that this would be disease-specific and cannot explain the lack of differences between disease- and control samples in terms of cell type proportions. We, therefore, reasoned that the observed disease-specific decrease in neuronal signal in the bulk RNA-seq data could be explained by: (i) a decrease in the extra-nuclear fraction of the transcriptome of neurons (i.e., loss of cytoplasmic and/or synaptic transcripts) and/or (ii) a decrease in the neuronal transcriptional output per neuron (i.e., an overall decrease in the number of nuclear transcripts per cell). To further explore this question, we compared the number of genes per barcode (i.e., per captured nucleus) between each of the three disease groups and controls, per cell type. This analysis indicated a general trend for decrease in the total number of expressed genes in 14/20 neuronal clusters in iPD and 14/20 in LRRK2-PD. Conversely, a trend for increase in the number of expressed genes was seen in 5/8 glial clusters in iPD and 7/8 in LRRK2-PD (Figure 2D). This trend was not observed in MSA. To formally test whether the number of expressed genes in neurons and glia was indeed altered in disease, we combined nuclei into three major clusters (excitatory neurons, inhibitory neurons, and glia, see Methods). As suggested by the exploratory analyses, iPD and LRRK2-PD exhibited a significant decrease in the number of expressed genes in both inhibitory and excitatory neurons, relative to glia, with the largest decrease exhibited in iPD. MSA exhibited only mild changes, which did not reach statistical significance (Supp. Table 2).

**Table 2.** Functional overrepresentation analysis in the α-synuclein pathology associated transcriptional signature. Significantly enriched GO terms in the differentially expressed genes associated with α-synuclein pathology in LRRK2-PD samples. Padj: Benjamini-Hochberg adjusted p; overlap: DEGs overlapping the gene list; size: size of the gene list; direction: direction of change of enriched DEGs.

| α-synuclein pathology in LRRK2-PD | | | | | |
| --- | --- | --- | --- | --- | --- |
| pathway | padj | overlap | size | direction |  |
| Mitochondrial ATP Synthesis Coupled Electron Transport | 4.01E-07 | 23 | 36 | upregulation | ex_06 |
| Aerobic Electron Transport Chain | 1.83E-06 | 22 | 35 | upregulation |  |
| Cellular Respiration | 8.19E-06 | 24 | 43 | upregulation |  |
| Oxidative Phosphorylation | 1.43E-04 | 21 | 38 | upregulation |  |
| Proton Motive Force-Driven ATP Synthesis | 1.53E-04 | 20 | 35 | upregulation |  |
| Mitochondrial Electron Transport, Cytochrome C To Oxygen | 8.85E-04 | 9 | 9 | upregulation |  |
| Proton Motive Force-Driven Mitochondrial ATP Synthesis | 1.30E-03 | 18 | 32 | upregulation |  |
| Chaperone-Mediated Protein Complex Assembly | 4.88E-02 | 8 | 9 | upregulation |  |
| Mitochondrial Membrane | 5.70E-09 | 69 | 205 | upregulation |  |
| Organelle Inner Membrane | 1.53E-08 | 54 | 144 | upregulation |  |
| Mitochondrial Inner Membrane | 1.63E-08 | 52 | 136 | upregulation |  |
| Mitochondrial Proton-Transporting ATP Synthase Complex | 5.82E-03 | 9 | 11 | upregulation |  |
| Mitochondrial Respiratory Chain Complex IV | 7.68E-03 | 8 | 9 | upregulation |  |
| Mitochondrial Respiratory Chain Complex I | 3.34E-02 | 12 | 21 | upregulation |  |
| Oxidoreduction-Driven Active Transmembrane Transporter Activity | 1.27E-06 | 19 | 28 | upregulation |  |
| Active Monoatomic Ion Transmembrane Transporter Activity | 1.44E-03 | 8 | 8 | upregulation |  |
| Ubiquitin Protein Ligase Binding | 4.31E-02 | 39 | 133 | upregulation |  |
| pathway | padj | overlap | size | direction |  |
| Cytoplasmic Translation | 8.40E-22 | 38 | 51 | upregulation | ex_11 |
| Peptide Biosynthetic Process | 2.53E-19 | 40 | 62 | upregulation |  |
| Macromolecule Biosynthetic Process | 5.28E-19 | 42 | 69 | upregulation |  |
| Translation | 4.89E-14 | 42 | 86 | upregulation |  |
| Gene Expression | 1.19E-13 | 50 | 120 | upregulation |  |
| Cytosolic Large Ribosomal Subunit | 7.00E-11 | 21 | 29 | upregulation |  |
| Vacuolar Proton-Transporting V-type ATPase Complex | 4.13E-02 | 8 | 12 | upregulation |  |
| Proton-Transporting V-type ATPase, V1 Domain | 8.25E-03 | 7 | 8 | upregulation |  |
| Polysomal Ribosome | 2.88E-02 | 10 | 18 | upregulation |  |
| Cytosolic Small Ribosomal Subunit | 1.08E-07 | 14 | 17 | upregulation |  |
| Ribosome | 7.24E-07 | 18 | 30 | upregulation |  |
| Small Ribosomal Subunit | 1.41E-08 | 15 | 18 | upregulation |  |
| pathway | padj | overlap | size | direction |  |
| Cytoplasmic Translation | 5.92E-04 | 14 | 36 | upregulation | in_12 |
| Peptide Biosynthetic Process | 1.53E-02 | 14 | 45 | upregulation |  |
| Translation | 3.93E-02 | 15 | 55 | upregulation |  |

### PD and MSA show distinct cell-specific expression profiles

Next, we assessed cell type-specific differential gene expression profiles. For each of the 28 cell type clusters, we calculated differential gene expression between each disease group and controls, accounting for the number of features in a droplet, age, sex, post-mortem interval (PMI) and disease status of the sample. Individuals were included in the model as a random effect to mitigate pseudoreplication bias^16,17^, greatly decreasing the p-value inflation (see Methods, Supp. Figure 1). We found a total of 272 differentially expressed genes (DEGs) in iPD, 54 in LRRK2-PD, and 125 in MSA, at FDR < 5% (complete results available as Supp. Data 1-3).

iPD and LRRK2-PD exhibited a widespread transcriptional downregulation in excitatory neurons (iPD: 31 upregulated and 115 downregulated DEGs; LRRK2-PD: 11 upregulated and 23 downregulated DEGs) and inhibitory neurons (iPD: 10 upregulated and 23 downregulated DEGs; LRRK2-PD: 1 upregulated and 3 downregulated DEGs; Figure 3A-B). The neuronal signature in iPD and LRRK2-PD was qualitatively similar, with 60% (23/38) of LRRK2-PD DEGs found also in the iPD group – in contrast to only 10% (4/38) of LRRK2-PD DEGs present in MSA (Figure 3G). Among all cell type clusters, the excitatory cluster ex_19 stood out in both idiopathic and monogenic forms of PD as the most affected cell type in the cortex (Figure 3A-B). The marker expression profile places ex_19 in the deep cortical layers (L6/L6b), characterized by the highest relative expression of *ADRA2A*, encoding the alpha 2A adrenoreceptor, across all excitatory subtypes (Figure 1E). Thus, these neurons appear to have a high density of adrenergic afferents.

**Figure 3.**
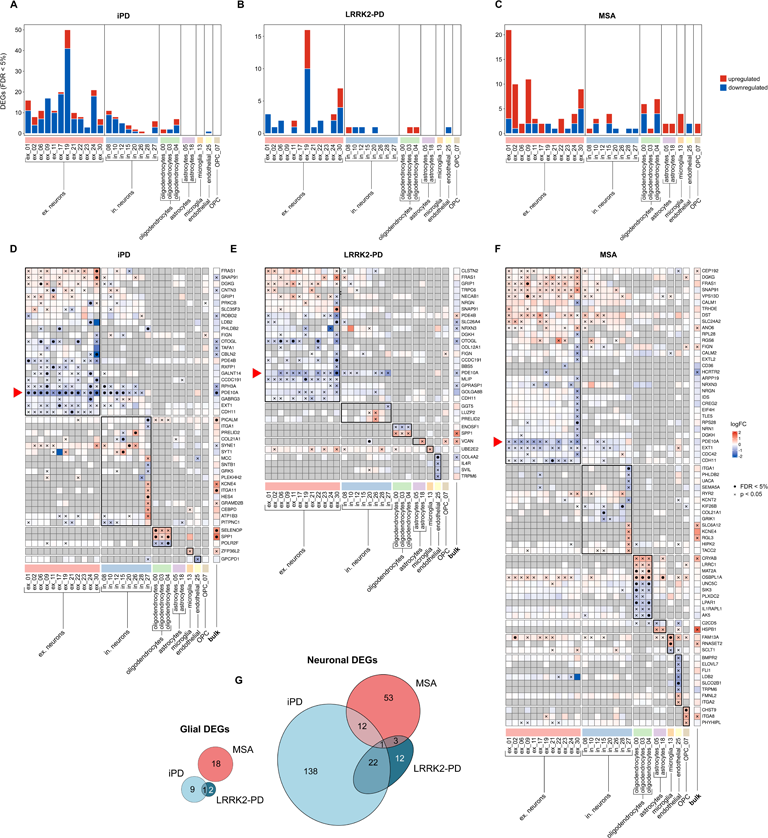
Differential gene expression within cell type clusters. **(A-C)** Number of differentially expressed genes at FDR < 5% (DEGs) in each cell type cluster. Each panel corresponds to the contrast between the disease (**A**: iPD, **B**: LRRK2-PD, **C**: MSA) and the healthy controls. **(D-F)** Heatmaps representing log fold-change in expression between diseases and controls in each cell type cluster (columns). Genes (rows) are only represented in the heatmaps if they exhibit a log2 fold change (logFC) below -0.5 or above 0.5 in at least one cluster. Nominal significance (p < 0.05) is represented with crosses, DEGs (FDR < 5%) are represented with black dots. Red triangles highlight the row corresponding to *PDE10A*. **(G)** Venn diagrams with neuronal and glial DEGs in the three diseases. Overlaps between contrasts represent common DEGs with concordant direction of change.

MSA showed a highly distinct transcriptional signature compared to iPD and LRRK2-PD. It exhibited a predominant upregulation of gene expression in excitatory neurons (42 upregulated and 17 downregulated DEGs), which was particularly evident in the most abundant excitatory subtypes of the outermost cortical layers (ex_01 and ex_02, layers L2/3, Figure 1E), but also manifest in ex_09, located deeper in the cortex. Contrarily to both PD groups, MSA did not display a pronounced dysregulation of the ex_19 neuronal subtypes (Figure 3C) and had a low concordance in neuronal DEGs with both iPD and LRRK2-PD (19% (13/69) of MSA DEGs found in iPD and 6% (4/69) present in LRRK2-PD; Figure 3G).

### *PDE10A* is downgulated in most PFC neurons in PD, LRRK2-PD and MSA

Most DEGs were highly cell type-specific, i.e., 81% (255/313) were unique to a cell type cluster. The remaining 19% of the DEGs occurred in 2-5 cell types, with one single striking exception: the gene *PDE10A* was found to be significantly downregulated in 14 clusters, comprising 10/12 excitatory neuronal types and 4/8 inhibitory neuronal types in iPD. Moreover, although *PDE10A* was only significantly downregulated in a single neuronal cluster in LRRK2-PD, it was consistently downregulated at the nominal level (uncorrected p < 0.05) in the majority of neuronal clusters in both LRRK2-PD (15/20 neuronal clusters) and MSA (12/20 neuronal clusters; Figure 3D-F). *PDE10A* encodes phosphodiesterase 10A, a protein known to be highly expressed in the human striatum^18^. Our data showed that *PDE10A* is abundantly expressed in all neuronal cell types and in endothelial cells of the human PFC, while its expression in glial cells is low (Figure 4A). To validate these findings at the protein level, we performed immunostaining on formalin-fixed paraffin-embedded sections of PFC from iPD patients (n=7) and controls (n=7). In line with the snRNA-seq data, the staining revealed a strong cytoplasmic expression of the PDE10A protein in PFC neurons and in endothelial cells (Figure 4C-E). Evaluation of the sections by two independent investigators who had been blinded to the disease group revealed a significantly weaker neuronal staining intensity in PD compared to controls (staining intensity score 0-3; iPD: 1.75 ± 0.59, controls: 2.57 ± 0.13, two-sided t-test: p = 0.009) with high interrater reliability (Cronbach’s alpha = 0.87; Figure 4B).

**Figure 4.**
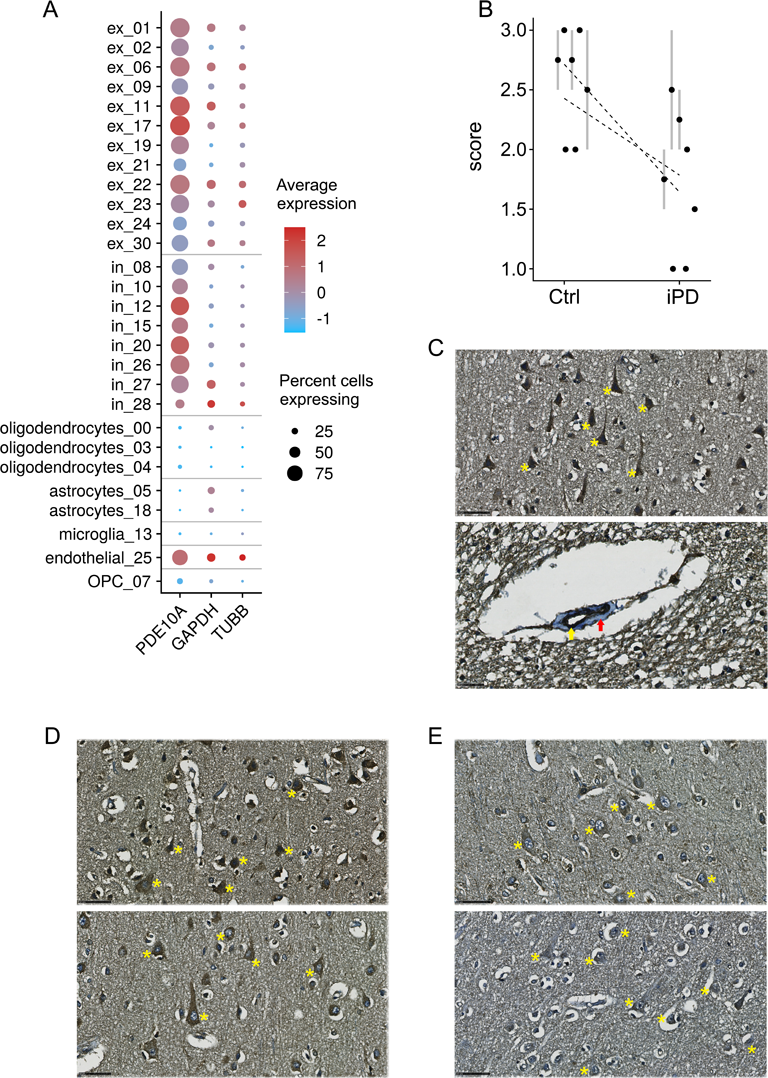
Expression of *PDE10A* in the PFC. **(A)** Dot plot shows the relative expression of the *PDE10A* transcript in each cell type cluster for the complete snRNA-seq dataset. Housekeeping genes *GAPDH* and *TUBB* are shown as a reference. **(B)** Quantification of immunohistochemistry images in iPD (n=7) and controls (n=7). The plot represents the mean score values (staining intensity scored from 0-3) between the two raters (black points) with grey bars spanning between the individual values o each rater when not in agreement (iPD: 1.75 ± 0.59, controls: 2.57 ± 0.13, two-sided t-test: p = 0.009). Dashed lines indicate linear regression for the scores for each rater. **(C-E)** Immunohistochemistry images for PDE10A. **(C)** PDE10A is highly expressed in neurons (upper panel) and endothelial cells (lower panel). Examples of neurons are indicated by yellow stars. The yellow and red arrows indicate the endothelial layer and muscle layer of the vessel, respectively. Upper panel: magnification x 500, scale bar: 50 µm. Lower panel: magnification x 800, scale bar: 25 µm. **(D-E)** Representative sections stained for PDE10A are shown from two controls **(D)** and two individuals with iPD **(E)**. Examples of neurons are indicated by yellow stars. Magnification x 500. Scale bar: 50 µm

### Disease-specific dysregulation of glial gene expression

Dysregulation of gene expression was also found within glial populations. Oligodendrocytes presented dysregulated expression in 9 genes in iPD (*ENOSF1*, *POLR2F*, *FOLH1*, *CD22*, *PCM1*, *SEMA3C*, *SPP1*, *NXX6-2* and *SELENOP*), 2 in LRRK2-PD (*FBXL17* and *SEMA3C*), and 8 in MSA (*AK5*, *SIK3*, *LPAR1*, *UNC5C*, *FUT8*, *EPM2AIP1*, *MAT2A* and *OSBPL1A*). With the exception of *SEMA3C*, which was upregulated in the oligodendrocytes_04 cluster in both iPD and LRRK2-PD, no DEGs overlapped between groups (Figure 5). In addition, MSA showed a significantly altered expression in 3 genes within the two astrocytic populations (*RAPGEF3*, *ABHD3* and *BNIP3L*), while no DEGs were found in the two PD forms. Of note, *BNIP3L* was consistently downregulated in all glial subtypes in MSA, albeit only nominally significant (p < 0.05). In the microglial population, a single DEG was detected in MSA (*RNASET2*) and two in the OPC cluster (*MAPT* and *CHST9*), whereas the two PD forms reported no significant differences in expression within these glial clusters. Finally, endothelial cells showed a single DEG for iPD (*DDX58*), and for LRRK2-PD (*COL4A2*), and two for MSA (*DDX60L* and *SLCO2B1;* Figure 5).

**Figure 5.**
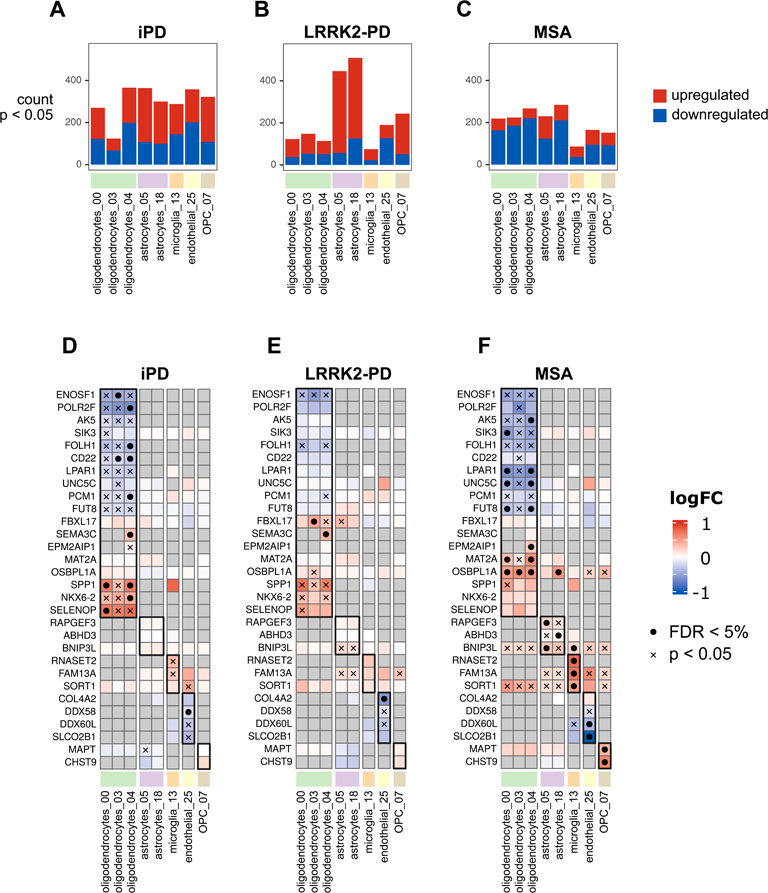
Differential gene expression within glial cell type clusters. **(A-C)** Number of differentially expressed genes at nominal p < 0.05 in each glial cell type cluster between disease groups and healthy controls. **(D-F)** Heatmaps represent log fold-change in expression between diseases and controls in each glial cell type cluster (columns). Genes (rows) are only represented in the heatmaps if they are significantly expressed (FDR < 5%) in at least one cell type cluster in at least one group. Black dots indicate statistical significance at FDR < 5%, crosses represent significance at the nominal level (p < 0.05).

### Pathway enrichment analysis highlights dysregulation of ribosomal pathways

Functional enrichment in DEGs was carried out by overrepresentation analysis of Gene Ontology (GO) pathways. The upregulated DEGs identified in iPD in the ex_19 and ex_21 clusters showed a significant enrichment in functions associated with protein translation, mostly due to the collective upregulation of ribosomal and associated transcripts. The same ribosomal-enriched signature was found in LRRK2-PD, albeit restricted to the ex_19 cluster, with the functional relevance of protein synthesis further highlighted by numerous non-ribosomal proteins (“RNA binding” GO term). In addition, iPD ex_19 was enriched in the GO term “MHC Class II Protein Complex Binding”, with 3/6 upregulated genes being heat shock protein transcripts (*HSP90AA1*, *HSPA8*, and *HSP90AB1*). Other overrepresented terms in upregulated DEGs in iPD were “Vesicle” (also in ex_19), “Calcium ion transmembrane import into cytosol” (in_27), and “Establishment of mitotic sister chromatid cohesion”, with the 5 components of the cohesin complex upregulated in ex_06 (Table 1, Supp. Data 4).

In contrast to PD, MSA exhibited a downregulation of ribosomal pathways. This was observed in neuronal cluster ex_30, which also showed a significant downregulation of nuclear encoded OXPHOS subunits. Additional overrepresented GO terms in MSA included “Microtubule binding” (upregulation in ex_01 and ex_06) and “GTPase binding” (upregulation in ex_02; Table 1, Supp. Data 4). Glial clusters did not exhibit significant functional overrepresentation in any of the α-synucleinopathies.

### Cell type-specific differential expression of genes with a known association to PD

A total of 69/91 PD GWAS-nominated protein-coding genes^19^ were detected in at least one cell type cluster. Among these, *VPS13C*, *IGSF9B* and *KCNIP3* were differentially expressed in iPD (FDR < 5%), each in one excitatory neuron cluster (ex_21, ex_19 and ex_09, respectively). Notably, *VPS13C* was overexpressed at a nominal level (p < 0.05) in all excitatory clusters except for ex_17 (Supp. Figure 2). In LRRK2-PD, no GWAS-nominated genes were among the DEGs (Supp. Figure 2). None of the known genes causing familial monogenic PD (*LRRK2*, *VPS35*, *PINK1*, PINK7, *PRKN* and *SNCA*) were differentially expressed in any of the PD forms (Supp. Figure 2). Overrepresentation analysis for the GWAS-nominated genes within each cell type cluster yielded no significant results at 5% FDR (Supp. Data 5).

### α-synuclein pathology is associated with a distinct neuronal gene expression signature

To discern cell-specific gene expression changes associated with α-synuclein pathology independently from the disease status, we compared LRRK2-PD individuals with Lewy pathology (n=3, Braak stage 4-5) to the LRRK2-PD individuals without Lewy pathology (n=4, Braak stage 0). Differential gene expression in each cell type cluster revealed a total of 754 DEGs (henceforth referred to as *“α-synuclein signature”*), comprising 589 downregulated and 165 upregulated DEGs. Notably, this was a far larger signal than those observed in the contrasts between diseases and controls (full results provided in Supp. Data 6). The α-synuclein signature was predominantly neuronal, with neuronal clusters collectively gathering 647/754 DEGs and only 111/754 DEGs detected in glial clusters. Furthermore, the largest part of the signal originated from the most abundant types of excitatory neurons, located in the outer cortical layers (ex_01 and ex_02, with 324 and 245 DEGs, respectively) and intermediate layers (ex_06, ex_11, with 125 and 161 DEGs, respectively; Figure 6A). This was in sharp contrast to the iPD- and LRRK2-PD-specific transcriptional signatures, where most of the transcriptional dysregulation was observed in deep cortical neurons (clusters ex_19 and ex_24; Figure 3A-B). In fact, we found that the similarity between Lewy pathology-associated and the disease-specific PD transcriptional signature was low, with only 5% (34/647) neuronal DEGs overlapping with iPD and 1% (6/647) with LRRK2-PD (Figure 6B). The overlap of neuronal Lewy pathology-associated DEGs with MSA was even lower, below 1% (4/647).

**Figure 6.**
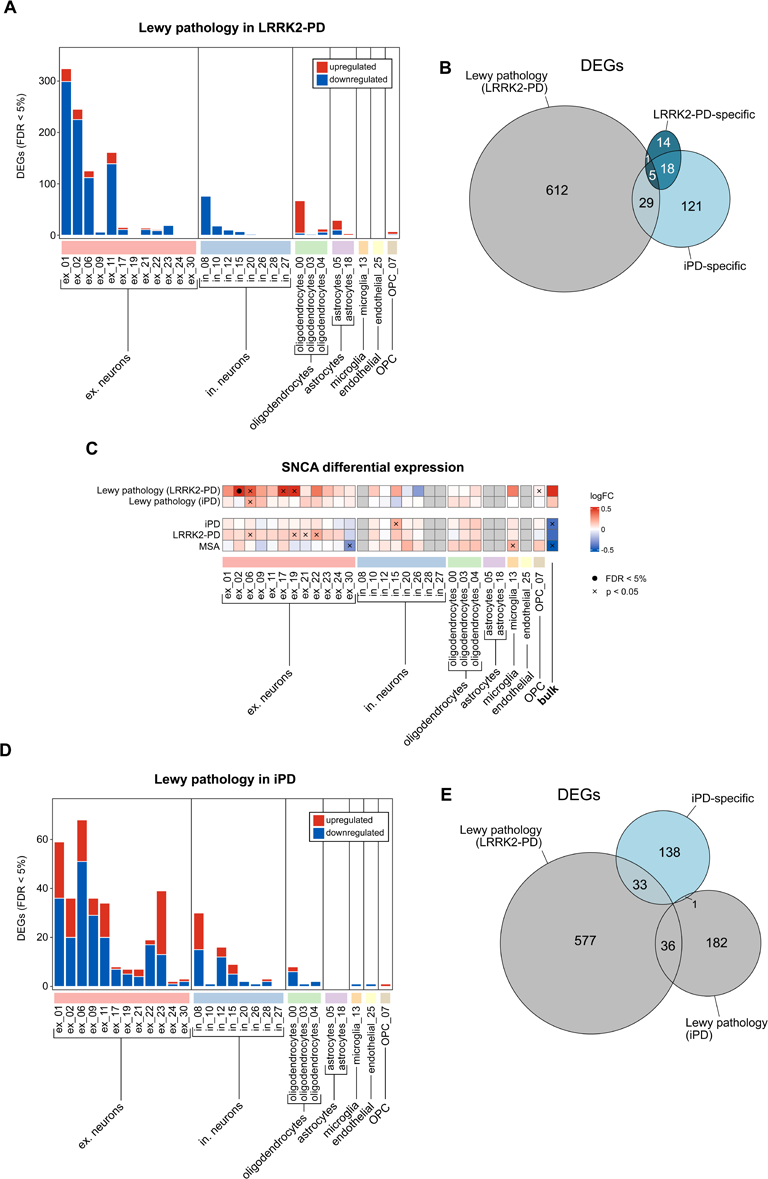
Differential gene expression associated with Lewy body pathology. **(A)** Number of differentially expressed genes (DEGs) at FDR < 5% in each cell type cluster between LRRK2-PD samples derived from brains without α-synuclein pathology (Braak 0) and LRRK2-PD samples from brains with α-synuclein pathology (Braak 4-5). **(B)** Venn diagram showing overlap between neuronal DEGs in α-synuclein pathology in LRRK2-PD samples (grey oval) and disease-specific neuronal DEGs – iPD *vs* controls (light-blue oval); LRRK2-PD *vs* controls (dark-blue oval). **C:** Heatmap representing log fold-change in expression of *SNCA* associated with α-synuclein pathology (within LRRK2-PD patients, top row; within iPD patients, second row) and between diseases and controls (3 lowest rows). Grey tiles correspond to expression levels below pre-filtering threshold in the cell type cluster. Black dots indicate FDR < 5%, crosses indicate nominal significance (p < 0.05). **(D)** number of DEGs associated with the degree of α-synuclein pathology in iPD samples (Braak range 4-6). **(E)** Venn diagram showing overlap in neuronal DEGs associated with Lewy pathology in LRRK2-PD samples, neuronal DEGs associated with Lewy pathology in iPD samples (grey ovals), and neuronal DEGs between iPD and controls (light-blue oval).

Despite the predominant downregulation signal observed in neurons, α-synuclein pathology was associated with a significant upregulation of the *SNCA* transcript in cluster ex_02 (Figure 6C). These are excitatory neurons located in the superficial cortical layers (L2/3) expressing *CBLN2* and *GNAL* genes (Figure 1E). While ex_02 showed the strongest signal and survived multiple testing correction, the upregulation of *SNCA* was consistent in all excitatory neuronal subtypes and nominally significant in ex_09, ex_17, and ex_19. Notably, altered *SNCA* expression was only seen in the α-synuclein transcriptional signature, and not observed in any of the disease-specific contrasts (Figure 6C). Other noteworthy DEGs associated with α-synuclein pathology included a neuronal upregulation of *HSP90AA1* and *HSP90AB1*, encoding heat shock proteins involved in cellular stress response^20^; *CALM*, encoding calmodulin, an important regulator of calcium signaling; and the *OPTN* and *SOD1* genes, encoding proteins involved in mitophagy and antioxidant response, respectively, mutations in which cause familial amyotrophic lateral sclerosis^21,22^. Additionally, we noted that oligodendrocytes exhibited an upregulation of *VPS13C*, involved in mitochondrial quality control, mutations in which cause recessive parkinsonism with diffuse Lewy body disease^23^ (Supp. Data 6).

Despite the general trend for downregulation, overrepresentation analysis of gene expression associated with α-synuclein pathology in LRRK2-PD individuals unveiled significant upregulation of processes related to mitochondrial respiration in clusters ex_06 and ex_11 (Table 2). This signal was driven by nuclear-encoded components of the respiratory chain. Additionally, we observed an upregulation of the ribosome and protein translation driven by ribosomal proteins, similar to that observed in the iPD- and LRRK2-PD disease-specific signatures, but originating from different neuronal subtypes (i.e., ex_11 and in_12). Finally, cluster ex_11 exhibited a significant upregulation of the GO term “Vacuolar proton-transporting V-type ATPase complex” (Supp. Data 7).

The common DEGs between the α-synuclein-associated signature in LRRK2-PD individuals and the disease-associated signature in iPD consisted of only 34 neuronal genes, of which 32 were downregulated. Despite not reaching statistical significance in the functional overrepresentation analysis, the downregulated signature was visibly composed of synaptic components (Supp. Data 8).

Finally, to identify an α-synuclein-associated signature in iPD, we carried out an analogous analysis in the iPD samples. Specifically, we assessed DEGs associated with the severity of Braak stage (range: 4-6). We identified a total of 226 DEGs (140 downregulated and 86 upregulated), whereof the majority (193/226) were dysregulated in excitatory neuronal clusters (Supp. Data 9). Notable among them was an upregulation of *BASP1*, encoding a synaptic protein involved in neurite outgrowth and plasma membrane organization^24^, which was found prominently upregulated in induced pluripotent stem cell-derived neurons carrying a pathogenic *SNCA* mutation and expressing α-synuclein pathology^25^.

A total of 37 DEGs were common between the α-synuclein-associated signature in LRRK2-PD and iPD. These included a downregulation, in several excitatory neuronal clusters, of *RPTOR*, encoding a component of the regulatory associated protein of the mammalian target of rapamycin (mTOR) complex 1 (mTORC1), which has a central role in regulating cell metabolism and autophagy^26^. No significant pathway enrichment was found in the overrepresentation analyses.

## Discussion

Here, we report cell type-specific changes in the expression of genes and molecular pathways, associated with each of iPD, LRRK2-PD and MSA, and also with the presence and severity of neuronal α-synuclein pathology. The vast majority of differentially expressed genes in the snRNA-seq dataset (296/318) were highly cell type-specific, and therefore not identified by bulk RNA-seq in the same samples. This is hardly surprising, given the high level of specialization and differential vulnerability to disease exhibited by different cell types in the nervous system, and highlights the importance of cell type-specific studies in neurodegeneration.

The transcriptomic signatures of iPD and LRRK2-PD were qualitatively similar, and highly distinct from that of MSA. While more genes were found to be significantly differentially expressed in iPD, most of these (201/252) were also nominally significant in LRRK2-PD, indicating that the difference is mostly due to the larger sample size and statistical power in the iPD group. The transcriptomic overlap between iPD and LRRK2-PD suggests overlapping disease-mechanisms. This is perhaps not entirely unexpected, given that LRRK2-PD is clinically and pathologically virtually indistinguishable from iPD – with the exception that not all LRRK2-PD individuals harbor α-synuclein pathology.

PD (i.e., either iPD or LRRK2-PD) exhibited a predominantly neuronal signature, with relatively few differentially expressed genes in glial cells. Strikingly, most of the differential expression signal in both iPD and LRRK2-PD was concentrated in the excitatory neuronal cluster ex_19. These deep cortical neurons (layers L6/L6b), express high levels of the alpha 2A adrenoreceptor and, therefore, likely receive adrenergic afferents from the noradrenergic neurons of the locus coeruleus^27^, a neuronal population typically degenerated in PD^28^. It is possible that the observed transcriptional dysregulation of these neurons is, at least in part, a result of their adrenergic denervation. This hypothesis is, however, contradicted by the fact that MSA does not recapitulate a preferential involvement of the ex_19 neurons, despite exhibiting severe degeneration of the locus coeruleus and loss of coeruleocortical afferents^29,30^.

Alpha 2 adrenoreceptors are decreased in the PFC of individuals with PD^31^, and alpha 2 adrenoreceptor agonists have been shown to ameliorate cognitive deficits in patients^32,33^. Interestingly, studies in nonhuman primates demonstrated that alpha 2 adrenoreceptor agonists exert these effects by acting mainly on the dorsolateral PFC^34^, which is the region our samples originated from. While the mechanisms underlying the selective involvement of the ex_19 neuronal cluster remain unknow, our findings suggest that aberrant function of these neurons plays a role in both iPD and LRRK2-PD. Whether this is related to impaired alpha 2 adrenergic transmission is unknown, and further research is warranted to understand the involvement of these neurons in PD, as well as the clinical correlates of their altered function.

Among the differentially expressed genes in our dataset, *PDE10A* stood out due to its strikingly consistent downregulation across nearly all cortical neuronal types, both excitatory and inhibitory, in iPD and LRRK2-PD. This observation was confirmed at the protein level by immunohistochemistry, showing decreased neuronal expression of the PDE10A protein in PD.

*PDE10A* encodes phosphodiesterase 10A, an enzyme playing an important role in signal transduction by regulating the intracellular levels of cyclic adenosine monophosphate (cAMP) and cyclic guanosine monophosphate (cGMP). PDE10A is highly expressed in human striatal medium spiny neurons, where it is involved in movement control. By hydrolyzing cAMP and/or cGMP, PDE10A inhibits dopamine D_1_ receptor signaling in the striatonigral pathway and enhances dopamine D_2_ receptor signaling in the striatopallidal pathway, thereby leading to an overall decrease of movement^18,35^. Using positron emission tomography, PDE10A has been shown to be decreased in the striatum of PD patients, possibly as a compensatory response to the loss of nigrostriatal dopaminergic input^18^. Moreover, PDE10A inhibitors improve parkinsonistic motor deficits in preclinical animal studies, via their action on the striatal pathways, and have been proposed as a potential dopamine-independent motor treatment for PD^35,36^. However, the physiological function and disease-relevance of PDE10A outside the striatum are to date unknown. We show that PDE10A mRNA and protein are highly expressed in neurons and endothelial cells of the human PFC, with much lower expression in glia. Moreover, PDE10A transcript and protein levels are dysregulated across most neuronal types of the PFC in PD (both iPD and LRRK2-PD), indicating that the implication of this protein in the pathogenesis of PD extends well beyond regulation of dopaminergic transmission in the striatum and motor control. Further investigation into the functional implications of *PDE10A* dysregulation in the PD cortex could provide valuable insights into molecular disease mechanisms and potential therapeutic targets.

The cell-specific transcriptomic signature of MSA was highly distinct from that of PD, both in terms of dysregulated genes and affected cell types. In addition to multiple differentially expressed genes in neurons, MSA exhibited a prominent oligodendroglial signal, in line with its glial pathology^1,2^. Moreover, MSA showed no predilection for the ex_19 neuronal cluster, and no significant dysregulation of the *PDE10A* gene.

Harnessing the variable presence of α-synuclein pathology in LRRK2-PD allowed us to discern cell-specific transcriptomic signatures associated with α-synuclein pathology in the human brain. Strikingly, this comprised a neuronal upregulation of the *SNCA* gene itself, as well as several genes involved in stress response to oxidative damage, calcium regulation, and mitochondrial function. At the pathway level, α-synuclein pathology was strongly associated with an upregulation of processes involved in mitochondrial respiration, including components of the respiratory chain itself. This phenomenon may represent a compensatory increase in the transcription of mitochondrial respiratory pathways in response to functional and/or quantitative respiratory deficiency, which has been shown to be associated with early forms of α-synuclein pathology in PD neurons^37^. However, since respiratory chain integrity has not been investigated in human brain tissue from LRRK2 disease^38^, it is currently unknown whether respiratory dysfunction occurs in these patients’ neurons.

iPD exhibited a generally weaker α-synuclein-associated signature compared to LRRK2-PD. This is likely due to the fact that all iPD individuals harbored advanced α-synuclein pathology, with a much smaller variation in severity (Braak stage 4-6) than that of LRRK2-PD (Braak stage 0-5). Thus, we believe that the analysis of LRRK2-PD is more sensitive and specific in this regard, and differential expression signatures should not be considered less relevant for not being present also in iPD. Nevertheless, altered expression in some genes was found to be associated with α-synuclein pathology in both LRRK2-PD and iPD, including neuronal downregulation of *RPTOR*. This finding suggests that a dysregulation of the mTORC1 pathway, which has a central role in regulating cell metabolism and autophagy^26^, may be involved in the pathogenesis of α-synucleinopathies. Aberrant mTOR/mTORC1 signaling has also been found in preclinical models of PD^39^. Moreover, mTOR/mTORC1 signaling can be modulated by a number of approved drugs, including the glucagon-like peptide 1 (GLP-1) receptor agonist exenatide, which are currently in trials for iPD, with encouraging results so far^40^.

Another noteworthy observation is that in iPD, the severity of α-synuclein pathology was associated with upregulation of *BASP1*. This gene encodes a synaptic protein involved in neurite outgrowth and plasma membrane organization^24^, and was found upregulated in induced pluripotent stem cell-derived neurons carrying a pathogenic *SNCA* mutation and expressing α-synuclein pathology^25^. Our findings indicate that *BASP1* dysregulation is associated with α-synuclein pathology in the brain of patients with iPD.

Finally, in addition to altered expression in specific genes and pathways, we identify a general trend for a disease-specific reduction in the overall neuronal transcriptional output. This intriguing phenomenon was more pronounced in inhibitory neurons and not encountered in glial cells. While its significance is currently unknown, it may reflect a general metabolic decline in dysfunctional neurons. In this context, inhibitory neurons may be more vulnerable due to their higher metabolic requirements^41^. Regardless of its biological significance, this phenomenon explains the observed discrepancy between bulk RNA-seq, indicating a decrease in the neuronal fraction and an increase in glial populations, and snRNA-seq data which indicated no change in cell composition. This has important methodological implications for the field of brain omics. Since current methods for estimating cell composition in bulk brain tissue transcriptomic data are based on the expression of cell type markers, they cannot distinguish between a reduction in the cell number and the transcriptional output of a particular cell type. Thus, bulk tissue transcriptomics-based reports of neuronal loss in PD may be driven, at least in part, by a reduction in overall neuronal transcriptional output. Another possible explanation may be the loss of synaptic input to the PFC from deeper regions, including the basal forebrain nuclei and locus coeruleus, which characterizes α-synucleinopathies. Since synapses also contain neuronal RNA, they contribute to the cell proportion estimates in bulk tissue, but do not influence snRNA data. These findings raise questions about the validity of using bulk RNA-seq data to assess cellular composition in complex neurodegenerative diseases and emphasize the importance of single-cell analyses to gain a more accurate understanding of cell-type-specific changes.

## Methods

### Sample cohort

Fresh-frozen prefrontal cortex (PFC) samples (Brodmann area 9) were obtained from a total of 46 individuals. The dataset comprised samples from n = 13 healthy individuals, n = 20 idiopathic PD patients, n = 7 patients with monogenic LRRK2-PDs (n = 6 p.G2019S and n = 1 p.R1441H), and n = 6 individuals with parkinsonian MSA. Brain samples were obtained from our brain bank for aging and neurodegeneration, including samples from the Norwegian Park West cohort^42^, the Netherland Brain Bank, the Barcelona Brain Bank, the London Neurodegenerative Diseases Brain Bank (Supp. Table 1). Variant calling on the aligned RNA-seq data on all patients revealed no known/predicted pathogenic mutations in genes implicated in Mendelian PD or other monogenic neurological disorders, with the exception of the LRRK2-PD samples, for which the expected variants could be confirmed. Healthy controls had no known neurological disease and were matched for age and sex. Individuals with PD fulfilled the National Institute of Neurological Disorders and Stroke^43^ and the UK Parkinson’s disease Society Brain Bank^44^ diagnostic criteria for the disease at their final visit. Ethical permission for this study was obtained from our regional ethics committee (REK 2017/2082, 2010/1700, 131.04). Informed consent was available from all individuals.

### Isolation of single nuclei, snRNA-seq library preparation and sequencing

Nuclei isolation procedure was performed as described in ^45^ with minor modifications. All procedures were performed on ice. For each sample, 50 - 85 mg of frozen human PFC were homogenized. Tissue was dissociated in 2 mL of homogenization buffer (320 mM sucrose, 5 mM CaCl_2_, 3 mM Mg(CH_3_COO)_2_, 10 mM Tris HCl pH 7.8, 0.1 mM EDTA pH 8.0, 0.1% IGEPAL CA-630, 1 mM DTT, and 0.4 U/μL RNase inhibitor (ThermoFisher Scientific) inside a 7 mL KIMBLE Dounce tissue grinder set (Merck), using 10 strokes with the loose pestle A followed by 10 strokes with the tight pestle B. Homogenized tissue was filtered through a 30 μm cell strainer and centrifuged at 1000 xg for 8 min at 4°C. The supernatant was discarded, and pellets were resuspended in 450 μL of 2% BSA (in PBS) containing 0.12 U/μL RNase inhibitor. Removal of debris from the nuclei suspension was achieved by density gradient centrifugation. Equal volumes (450 μL each) of nuclei suspension and working solution (50% OptiPrep density gradient medium (Merck), 5 mM CaCl_2_, 3 mM Mg(CH_3_COO)_2_, 10 mM Tris HCl pH 7.8, 0.1 mM EDTA pH 8.0, and 1 mM DTT) were mixed by pipetting until the solution appeared homogeneous. The density gradient was prepared by carefully layering 750 μL of 30% OptiPrep Solution containing134 mM sucrose, 5 mM CaCl_2_, 3 mM Mg(CH_3_COO)_2_, 10 mM Tris HCl pH 7.8, 0.1 mM EDTA pH 8.0, 1 mM DTT, 0.04% IGEPAL CA-630, and 0.17 U/μL RNase inhibitor) on top of 300 μL of 40% OptiPrep Solution containing 96 mM sucrose, 5 mM CaCl_2_, 3 mM Mg(CH_3_COO)_2_, 10 mM Tris HCl pH 7.8, 0.1 mM EDTA pH 8.0, 1 mM DTT, 0.03% IGEPAL CA-630, and 0.12 U/μL RNase inhibitor) inside a 2 mL Sorenson Dolphin microcentrifuge tube (Merck). 800 µl of the mixture containing the nuclei were slowly pipetted onto the top of the OptiPrep density gradient and samples were centrifuged at 10,000 xg and 4°C for 5 min using a fixed angle rotor (FA-45-24-11-Kit, Eppendorf). Nuclei were recovered by withdrawing a volume of 200 μL from the 30 – 40% interface of the OptiPrep density gradient and transfer to a new 2 mL Sorenson Dolphin microcentrifuge tube (Merck). Nuclei were washed with 2 ml 2% BSA (in PBS) containing 0.12 U/μL RNase inhibitor and then pelleted by centrifugation at 300 xg and 4°C for 3 min using a swing-bucket rotor (S-24-11-AT, Eppendorf). This step was repeatedthree times. The nuclear pellet was then resuspended in a minimum volume of 100 μL of 2% BSA (in PBS) containing 0.12 U/μL RNase inhibitor and carefully mixed by pipetting 20 times on ice. The nuclei suspension was filtered through a 40 μm FlowMe cell strainer (Merck) to remove any remaining nuclei aggregates. Nuclei were stained with Trypan blue and counted manually on a hemocytometer.

A total of 10,000 nuclei per sample were targeted for droplet-based snRNA sequencing. cDNA synthesis and DNA library preparation were carried out using the Chromium Next GEM Single Cell 3′ Reagent Kit v3.1 Dual Index (10x Genomics) according to the manufacturer’s protocol. DNA Libraries were pooled and run using paired-end (300 million read pairs per sample) sequencing on the NovaSeq 6000 platform (Illumina) at Novogene (Cambridge).

### Bulk tissue RNA-seq library preparation and sequencing

Total RNA was extracted from PFC tissue homogenate for all samples using RNeasy plus mini kit (Qiagen) with on-column DNase treatment according to the manufacturer’s protocol. The final elution was made in 65 μl of dH_2_O. The concentration and integrity of the total RNA was determined by the Ribogreen assay (Thermo Fisher Scientific), and Fragment Analyzer (Advanced Analytical), respectively, and 500 ng of total RNA was used for downstream RNA-seq applications. First, rRNA was removed using Ribo-Zero™ Gold (Epidemiology) kit (Illumina, San Diego, CA) using the manufacturer’s recommended protocol. Two samples did not pass QC and were not sequenced (Supp. Table 1). Immediately after rRNA removal, the RNA was fragmented and primed for the first strand synthesis using the NEBNext First Strand synthesis module (New England BioLabs). Directional second strand synthesis was performed using NEBNext Ultra Directional second strand synthesis kit. Then, the samples were taken into standard library preparation protocol using NEBNext® DNA Library Prep Master Mix Set for Illumina® with slight modifications. Briefly, end-repair was done followed by poly(A) addition and custom adapter ligation. Post-ligated materials were individually barcoded with unique in-house Genomic Services Lab (GSL) primers and amplified through 12 cycles of PCR. Library quantity was assessed by the Picogreen Assay (Thermo Fisher Scientific), and the library quality was estimated by utilizing a DNA High Sense chip on a Caliper Gx (Perkin Elmer). Accurate quantification of the final libraries for sequencing applications was determined using the qPCR-based KAPA Biosystems Library Quantification kit (Kapa Biosystems, Inc.). Each library was diluted to a final concentration of 12.5 nM and pooled equimolar prior to clustering. One hundred twenty-five bp Paired-End (PE) sequencing was performed on an Illumina HiSeq2500 sequencer (Illumina).

### Single-nucleus RNA-seq analyses

snRNA-seq raw FASTQ files were aligned to the human genome (GRCh38) using CellRanger v.6.1.2 (10x Genomics) accounting also for intronic reads to include all nuclear transcripts (include-introns option) and excluding secondary mappings (*nosecondary* option). To estimate intronic:exonic ratio, an additional alignment was carried only accounting for reads mapped to exonic reads. Version 35 of the ENCODE transcriptome assembly (Ensembl 101) was used.

As it has been previously pointed out^46^, snRNA-seq of brain tissue suffers from an enrichment of neuronal transcripts in the ambient RNA which is captured together with the nuclei. To minimize the confounding effect of non-nuclear ambient RNA and discard empty droplets, we employed CellBender^47^ on the raw data prior to importing the counts into R, as it has been shown to significantly ameliorate this problem^46^. Raw counts for each sample obtained with CellBender were imported into R excluding droplets with less than 500 features, less than 1,000 counts, more than 3% mitochondrial reads or more than 2% ribosomal reads, resulting in a total of 403,146 non-empty droplets. In order to filter droplets for downstream analyses, we used a two-pass filtering strategy. In the first pass, following sample-specific normalization, automatic feature selection was carried out using Seurat^48^ v4.3.0.1 (*NormalizeData*, *FindVariableFeatures*) with default parameters. This was followed by integration using *SelectIntegrationFeatures* to scale and obtain principal components (PC) of gene expression (*ScaleData* and *RunPCA*). Using the first 50 PCs, all n = 46 samples were integrated using reciprocal PC analysis with automatically detected integration anchors (*FindIntegrationAnchors*) and using the n = 13 controls to build a reference (*IntegrateData*). Clustering was carried out using *FindNeighbors* and *FindClusters* with the first 30 PCs and 0.1 resolution, respectively, resulting in 22 clusters. Doublets were estimated independently for each individual sample using doubletFinder^49^ with pK = 0.01 and pN = 0.25. We noted that cluster 17 was highly enriched in doublets (more than half of the barcodes were flagged) and hence was removed for downstream analyses. Two other small clusters were also removed due to a high content of non-nuclear RNA (low *MALAT1* expression and high exonic ratio, cluster 20) and for having virtually all barcodes from a single sample (cluster 21; Supp. Figure 3). Barcodes were further filtered out if they fulfilled any of the following conditions: (i) above the 90 percentile in UMI counts, (ii) above the 90 percentile in number of genes, (iii) identified as doublet. The resulting dataset (340,644 barcodes) was then re-clustered for a second filtering pass using a higher resolution (res = 0.5) after scaling. In this step, the expression of cell type markers was used to guide a preliminary annotation of the clusters using SATB2 and SLC17A7 for excitatory neurons, GAD1 and SLC6A1 for inhibitory neurons, MOG and MOBP for oligodendrocytes, GFAP and AQP4 for astrocytes, CSF1R and CSF3R for microglia, CLDN5 and SLC2A1 for endothelial cells OLIG1, and VCAN and OLIG1 for oligodendrocyte precursor cells (OPCs). The marker expression allowed a straightforward identification of clusters composed of doublets and undefined populations of cells that were henceforth removed from the analyses. The resulting 28 final clusters (12 clusters of excitatory neurons, 8 of inhibitory neurons, 3 of oligodendrocytes, 2 of astrocytes, 1 microglial cluster, 1 OPC cluster and 1 endothelial cluster) were trimmed by removing barcodes which had zero UMI counts for either of the two respective major cell type markers, resulting in a final dataset of 299,582 barcodes (121,701 excitatory neurons, 44,711 inhibitory neurons, 83,302 oligodendrocytes, 26,178 astrocytes, 14,755 OPCs, 6,897 microglia, and 2,038 endothelial nuclei; Figure 1).

Cell type markers specific for each subcluster were identified using MAST^17^ on the log-transformed count data with the number of features per droplet and individual as fixed effects. Only protein-coding genes that were expressed in at least 25% of the droplets in at least one subcluster were used, and mitochondrial-encoded transcripts were removed. To alleviate computational needs, only a random subset of 100,000 out of the 299,582 nuclei were used to find markers for each cluster, proportionally to the specific cluster sizes. Additionally, barcodes were not considered if they were marked as outliers in terms of total counts, number of features, or number of mitochondrial-expressed transcripts within each cluster. Marking of outliers was computed with a modification to Tukey fences for skewed distributions^50^.

The general classification of neuronal subclusters as excitatory or inhibitory was conspicuously confirmed by the expression of *SATB2*, *SLC17A7*, *GAD1*, *SLC6A1* (Figure 1C). Based on markers described previously^51^, subclusters of excitatory neurons were classified according to cortical layer source (Figure 1E), and inhibitory neurons according to their cortical layer locations, class of interneuron and developmental subpallial origin (Figure 1F).

### Bulk tissue RNA-seq analyses

Bulk RNA-seq was carried out for neighboring tissues from the same individuals. A total of 44/46 samples succeeded library preparation (2 control samples failed QC, see Supp. Table 1). We used Salmon^52^ v1.3.0 to quantify the abundance at the transcript level with the fragment-level GC bias correction option (gcBias) and the appropriate option for the library type (ISR) against the GENCODE annotation release 35. Transcript-level quantification was collapsed onto gene-level quantification using the tximport^53^ R package version 1.28.0. We restricted the analyses to protein-coding genes in autosomal chromosomes.

### Cell type quantification

Cell type quantification was carried out using marker gene profiles (MGPs)^14^. MGPs are based on the across-sample relative expression of cell type markers, estimating cellularity from a volumetric perspective (i.e., proportion of the transcriptomic library originating from each cell type) rather than estimating the number of cells (i.e., the number of nuclei). The MGP estimates are thus influenced by a combination of (a) relative cell type abundance (i.e., number of cells) and (b) relative cell type-specific transcriptional output (i.e., cell types with more RNA content will result in larger estimates). MGPs were calculated for the bulk RNA-seq expression and for the snRNA-seq pseudobulk expression (per-sample aggregated reads of all filtered barcodes). Additionally, we considered the proportion of nuclei labelled in the snRNA-seq dataset to each major cortical cell type as an alternative cell type estimate. Prior to the calculation of MGPs, count data was normalized using a variance stabilizing transformation (*vst* function from the DESeq2^54^ R package v1.40.2). MGPs were calculated using the cortical markers from the NeuroEspresso database^14^ with human orthologs defined using the Homologene^55^ R package v1.4.68 as previously described^8^.

To assess differences in cortical cell type proportions between diseases and controls, we employed a linear regression for the MGP-based estimates accounting for age, sex, PMI and disease status. For the estimates based on the number of nuclei, counts were modelled using a generalized linear regression using a negative binomial distribution accounting for age, sex and PMI.

### Analysis of the effect of demographic variables in the cortical transcriptome

We correlated demographic variables (age, sex and PMI) with MGP-based cell type estimates in the bulk RNA-seq samples to reveal that age and PMI were strongly associated with the MGP-based cell type estimates (Supp. Figure 4). In particular, longer PMIs were associated with a relative decrease in the neuronal signal, in agreement with previous observations^56^. To further characterize these sources of variation, we modelled (i) the number of detected genes per droplet and (ii) the proportion of nuclei in the snRNA-seq dataset as a function of the demographic variables independently for each of the 28 cell type clusters. As expected, we observed a strong, consistent decrease in the transcriptional volume with longer PMIs in all neuronal clusters, a trend that was absent in glial cells (Supp. Figure 5A). The proportion of captured neuronal nuclei, however, did not show a consistent variation across neuronal subtypes (Supp. Figure 5B). Age was, if anything, positively associated with transcriptional output, although only in some clusters and with no marked differences between neuronal and glial populations. These results highlighted the importance of controlling for demographic covariates in downstream analyses.

In order to test for a relative decrease in the transcriptional volume in neurons, we used the number of detected genes per droplet as a proxy. We defined three “major clusters” (excitatory neurons, inhibitory neurons and glia) and modelled the number of genes per droplet as a function of age, sex, PMI and disease status, adding interaction terms for disease status × major cell type and accounting for differences between cell type clusters (nested within the “major clusters”) with a random intercept, while the cell type-specific effect of PMI was accounted for with a random slope and intercept. The resulting R formula was

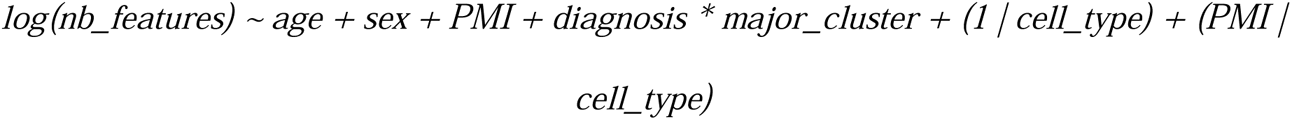

Age and PMI were scaled and centered, while the other categorical variables were modelled as dummy variables.

### Differential gene expression

Differential gene expression (DGE) analysis in the snRNA-seq dataset between each diagnosis (iPD, LRRK2-PD, MSA) and controls was carried out independently for each cluster by fitting a two-part Hurdle model for each gene as implemented in the MAST R package^17^ v1.24.1. The regressions were limited to the genes that were detected (i.e., with counts > 0) in 25% of the barcodes in the cluster. Read counts were normalized by the barcode library size and log2 transformed. In addition to sex and age, post-mortem interval (PMI) was used as a covariate since it showed a significant negative association with the proportion of intronic reads (i.e., longer PMI was associated with lower proportions of intronic reads). To account for the complex interaction between demographic variables (PMI and age) and cell type identity in modulating transcript levels (Supp. Figure 5), DGE analyses were always carried out independently for each of the 28 identified cell type clusters. In addition, and as suggested previously^17^, the proportion of genes detected in each cell (*prop_features*) was accounted for in the model as a fixed effect. Finally, to address the highly inflated type 1 error rates due to pseudoreplication, we included a random effect for individual^16^. The resulting R formula employed to model the expression of a gene *y* within a given cell type cluster was

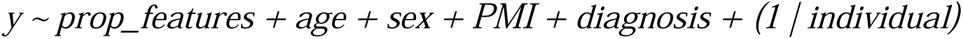

Adjusting for individual as a random effect resulted in a massive reduction of the p-value inflation and a decrease in the DGE of cell type markers within clusters when testing for differences between disease and controls (Supp. Figure 1). Assessment of statistical significance in DGE between diseases and controls was carried out using likelihood-ratio tests with the *lrTest* function from the MAST R package^17^ v1.24.1. The same package was used to calculate log fold-change estimates. P-values were corrected for multiple testing using the Benjamini-Hochberg false discovery rate (FDR) for each cluster.

To characterize the transcriptional signature associated with brain α-synuclein deposition, we first carried out DGE analysis between the LRRK2-PD samples without alpha-synuclein deposition (Braak stage 0; n=4) and LRRK2-PD samples with Braak stages 4-5 (n=3). DGE analysis was carried out without accounting for sex since all Braak stages 4 and 5 samples were from female individuals. The resulting R formula used was

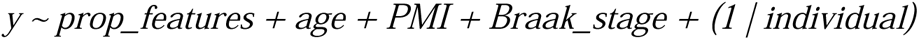

with *Braak_stage* taking the values 0 (for Braak stage 0) or 1 (for Braak stage 4 or 5). In addition, we carried out an independent DGE analysis using only iPD samples employing the same model design (i.e., accounting for Braak staging). In this case, sex was also accounted for in the formula and the *Braak_stage* variable taking values 4-6. For both analyses (LRRK2-PD subset and iPD subset), assessment of DGE was carried out analogously to the case-control contrast, but testing for the effect of Braak stage instead.

The subset of GWAS-nominated genes was initially obtained from ^19^, and non protein-coding transcripts filtered out resulting in a total of 91 genes. Only protein-coding genes were DGE from the bulk tissue RNA-seq data was performed using the DESeq2 R package version 1.40.2^54^ with default parameters and as previously described^8^. The generalized linear model was fit for each gene accounting for sex, age, PMI and disease status.

### Gene set enrichment analyses

In order to test for enrichment of functional terms within the DEG lists, we ran overrepresentation analyses using the fora() function from the R fgsea package^57^ v1.26 with the entire Gene Ontology database obtained from the enrichR database^58^. For a given cell type cluster, we used the nominally significant genes as input and the expressed genes in the cluster as background to minimize trivial enrichment in cell type markers. Overrepresentation analyses were carried out independently for up- and downregulated genes. When a list of DEGs consisted of the intersection between two contrasts, the overlapping input gene lists were restricted to DEGs from the same cell type clusters and with concordant directions of change. Multiple testing correction was calculated using Benjamini-Hochberg false discovery rate.

### Immunohistochemistry and image analyses

Immunohistochemistry analysis was performed on 3.5 µm thick formalin-fixed, paraffin-embedded PFC sections from 14 individuals (iPD n=7; controls n=7) used also in the snRNAseq analyses. Deparaffinization and antigen retrieval were performed in the low pH EnVision FLEX Target Retrieval Solution at 98 °C for 24 minutes in the DAKO PT link from Agilent. Sections were quickly rinsed with TBS Automation Wash buffer (Biocare, TWB945M) and then blocked with hydrogen peroxide (Peroxidased 1, Biocare, PX968M) for 10 min. After 5 min washing in TBS, sections were incubated in a humidity chamber for 1h at room temperature with rabbit anti-PDE10A (Abcam, ab151454) diluted 1:1000 in Da Vinci Green Diluent (Biocare, PD900M). Sections were washed for 5 min in TBS and then incubated with HRP-Polymer (Biocare, MRH534L) for 30 min. After two rounds of 5 min washing in TBS, DAB chromogen (Biocare, DB801) was applied for 4 minutes. After the visualization step, sections were washed in tap water for 30 seconds, counterstained with Tachas’s Hematoxylin (Biocare, NM-HEM-M) for 3 minutes and washed again in tap water for 5 min. Finally, sections were dehydrated by immersion in a series of graded alcohol solutions (from 75% EtOH to 100% EtOH) and Xylene and covered with glass coverslips using Pertex mounting medium (Histolab, 00811). Slides were digitally scanned using the NanoZoomer XR (Hamamatsu). Neuronal staining intensity was assessed visually, using the NDP.view2plus version(v) 2.7.25 software (Hamamatsu), using a scale of 0-3, with 0 indicating a lack of staining and 3 indicating an intense dark brown staining. Each section was assessed by two independent raters, blinded to the disease group. Interrater reliability was tested using Cronbach’s alpha coefficient. Since reliability was high, the mean value from both raters was used in downstream analyses. Normality was tested using the Shapiro Wilk test and homogeneity of variances was tested using the Levene’s test. Statistical group comparison was performed using a two-sided t-test.

## Supporting information

Supplementary Figures and captions for Supplementary Material

## Data Availability

All data produced in the present study are available upon reasonable request to the authors

## Acknowledgements

We are deeply grateful to the study participants and their families involved in the study for their unique contribution. Furthermore, we would like to thank the entire Park West study consortium for their diligent efforts characterizing and following the Park West cohort; the Neurological Tissue Bank of the IDIBAPS-Hospital Clinic for providing data and samples; the Netherlands Brain Bank; the London Neurodegenerative Diseases Brain Bank; Dr. Romain Guitton, Dr. Kristoffer Haugarvoll and Janani Sundaresan for helping dissect and prepare the brain tissue; Gry Hilde Nilsen for technical support.

## Availability of data and materials

The datasets and code required to reproduce the results of these analyses are available in the Neuromics Group repository, https://git.app.uib.no/neuromics/

## Authors’ contributions

GSN: participated in the study conception and design, designed and performed the analyses, and drafted the manuscript. MGC: establish the methodology for- and performed nuclei isolation and library construction and quality control on the 10X Genomics platform, and contributed to drafting the manuscript. SM: performed nuclei isolation and library construction on the 10X Genomics platform and contributed to drafting the manuscript. AR: performed and interpreted the immunohistochemistry experiments and contributed to drafting the manuscript. OS: contributed to the immunohistochemistry experiments. CD: contributed to the study design, establishing of the methodology, drafting of the manuscript, and performed nuclei isolation and library construction on the 10X Genomics platform. GA and OBT: contributed biological material. CT: conceived, designed and directed the study, contributed to data analyses and interpretation, drafted the manuscript, and acquired funding for the study. All authors have read and approved the manuscript.

## Ethics declarations

Ethics approval for these studies was obtained from our regional ethics committee (REK 2017/2082, 2010/1700, 131.04). Consent for publication was provided by all participants. The authors declare that they have no competing interests.

