## Supplementary Figures and captions for Supplementary Material for "Single-nucleus transcriptomics reveals disease- and pathology-specific signatures in α-synucleinopathies"

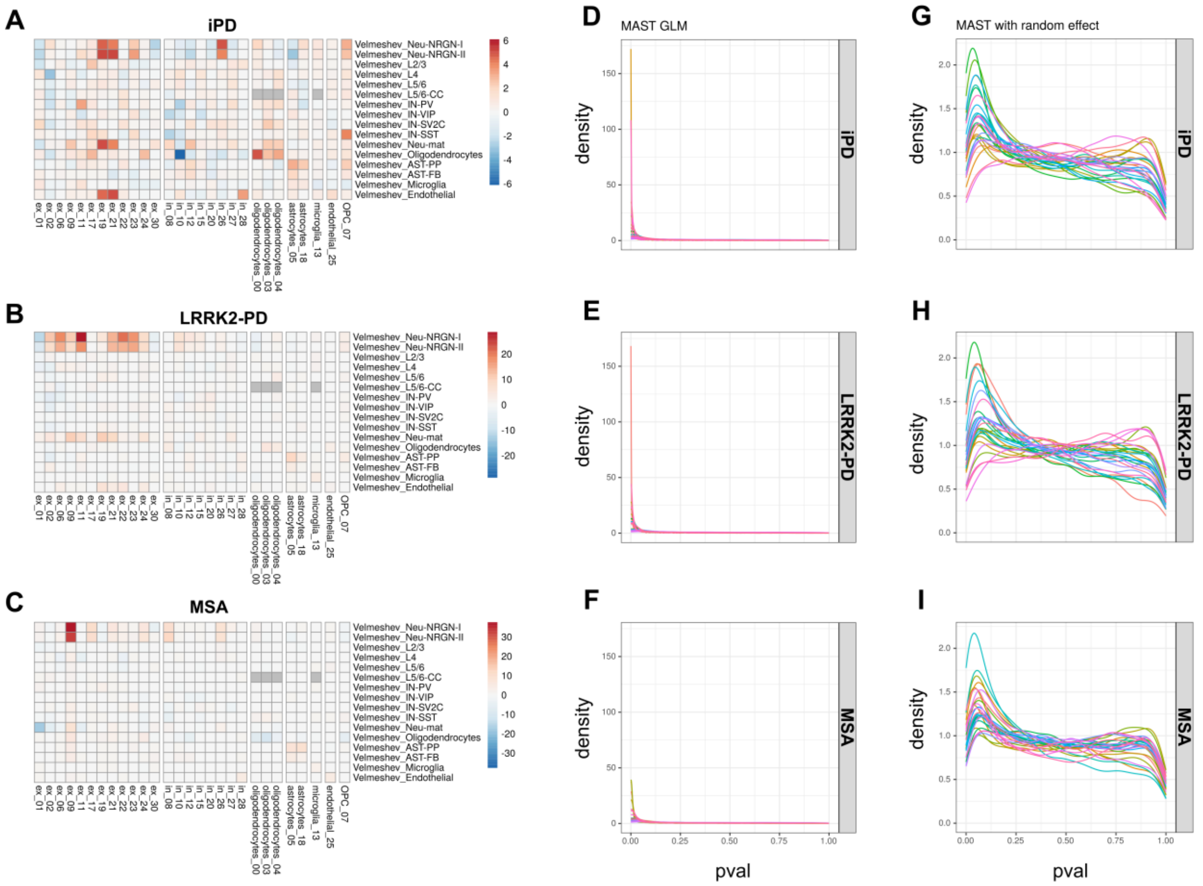


**Supplementary Figure 1**

**Influence of accounting for individual as random effect. A-C:** Gene set enrichment analysis carried out independently for the two alternative models (with or without individual as random effect) on a set of cell type specific pathways[1]. The difference of the -log10(pvalue) for the enrichment tests is represented in the heatmaps (red/positive values: decrease in the significance for the pathway; blue/negative values:increase in the significance). **D-I:** Distribution of the nominal p-values for the DGE tests carried out without the random effect (D-F) and with the random effect (G-I) (Y-axes in the density distribution plots are not aligned).

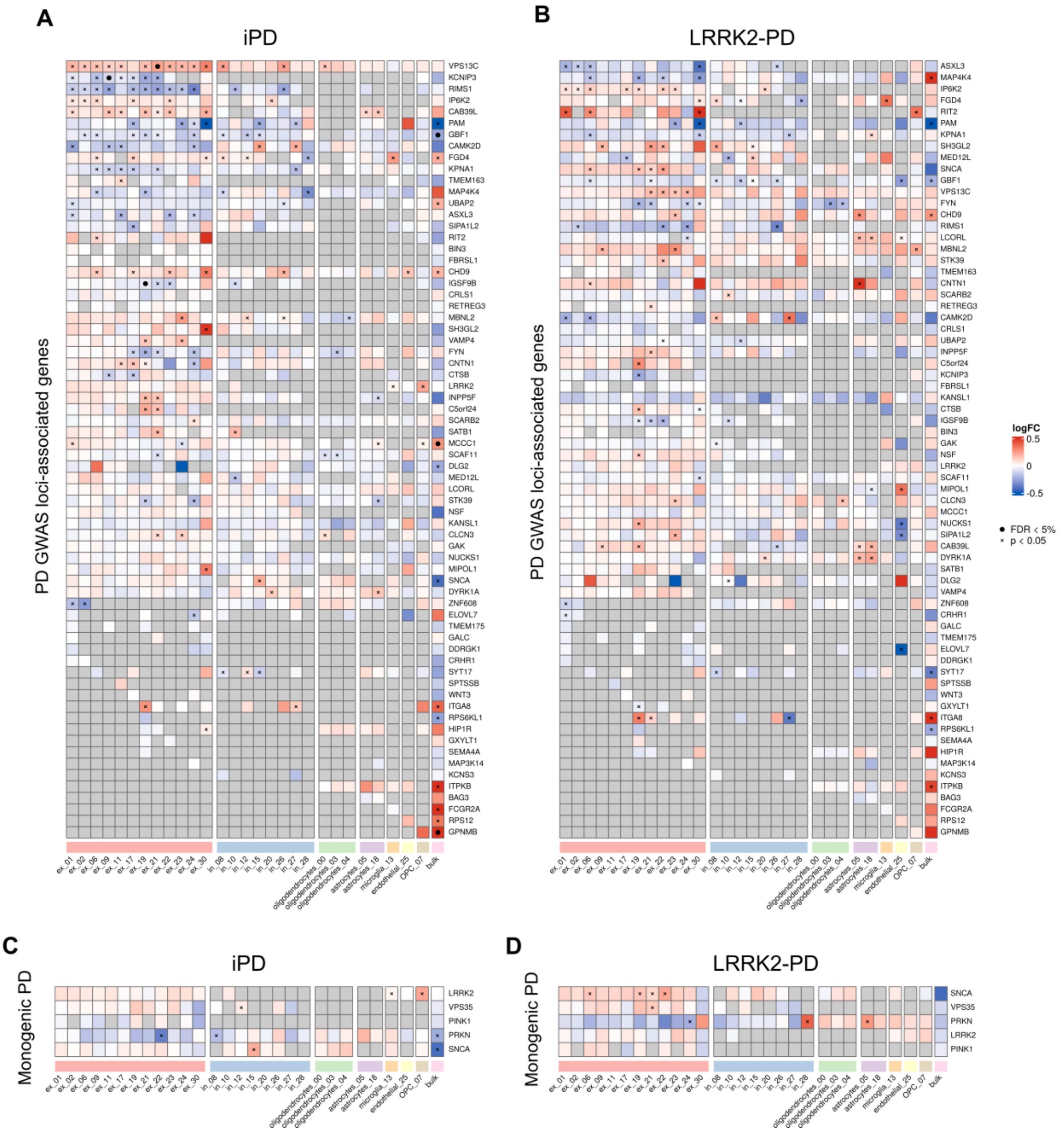


**Supplementary Figure 2**

**Differential expression of iPD-associated genes.** Heatmaps representing log2 fold-change in expression between iPD **(A, C)** and LRRK2-PD **(B, D)** against controls in each cell type cluster (columns). Genes (rows) correspond to the nearest genes to the PD GWAS-identified loci **(A, B)** and genes causing monogenic PD **(C, D)**. Black dots indicate FDR < 5%, crosses indicate nominal significance ($p < 0.05$).

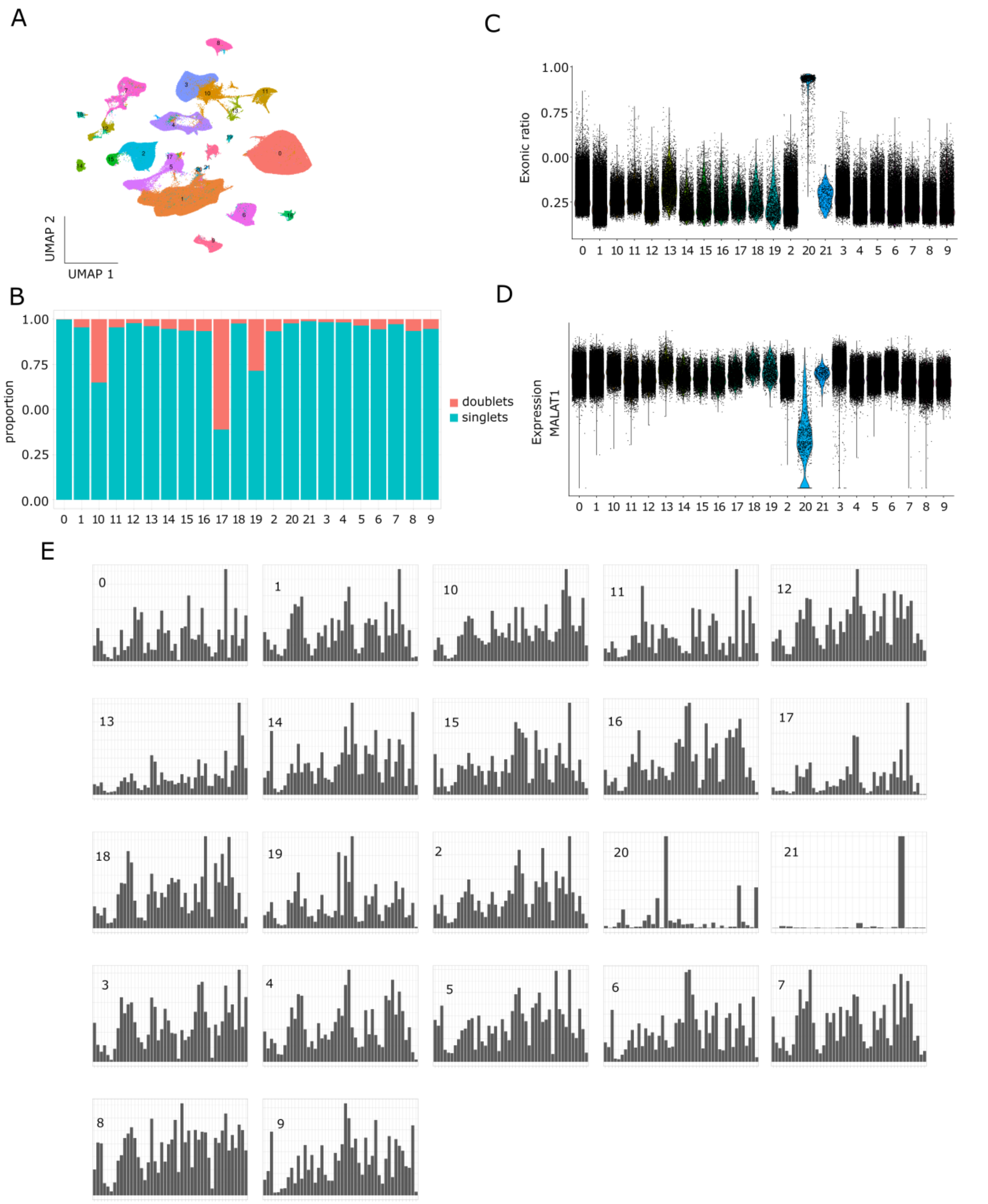


**Supplementary Figure 3**

**Prefiltering of snRNA-seq dataset.** **(A)** UMAP embeddings for the expression of 403,146 single nuclei from CellBender (barcodes with less than 500 features, less than 1,000 counts, more than 3% mitochondrial reads or more than 2% ribosomal reads have been excluded). Clustering was carried out at 0.1 resolution using Seurat. **(B)** Relative proportions of singlets and doublets as estimated by doubletFinder. Cluster 17 presents more than half of the droplets predicted as doublets. **(C)** Exonic ratio per droplet (i.e., median exonic counts / median exonic+intronic). Cluster 20 shows as an outlier. **(D)** Expression levels of *MALAT1* per cluster, also nominating cluster 20 as an outlier. **(E)**

Number of barcodes per cluster (panels) per sample individual (bars). Cluster 21 was composed almost entirely of barcodes from the same sample.

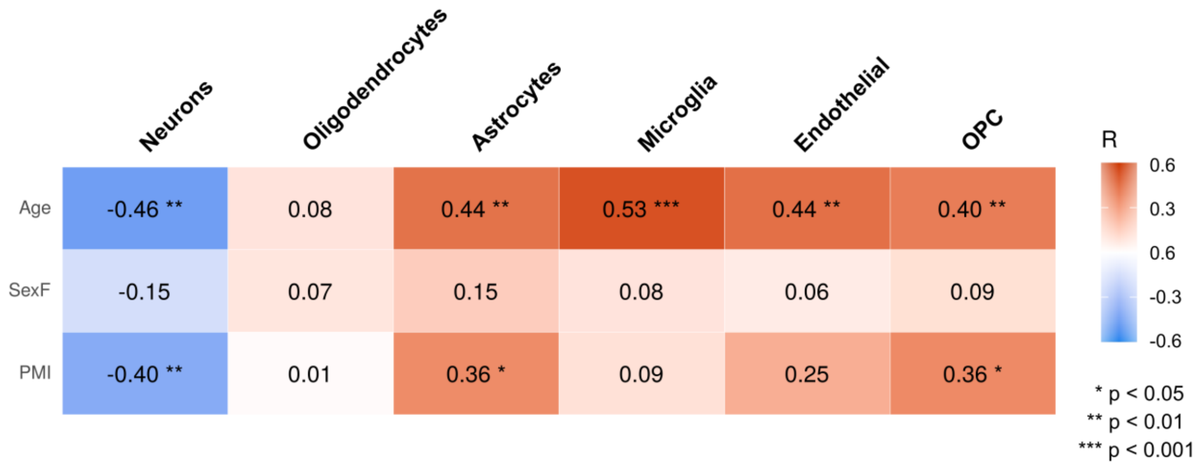


**Supplementary Figure 4**

**Correlation between demographic variables and cell type composition in bulk tissue**. The table shows the Pearson correlation coefficients between age, sex and PMI with the MGP-based cell type estimates calculated for the bulk RNA-seq samples.

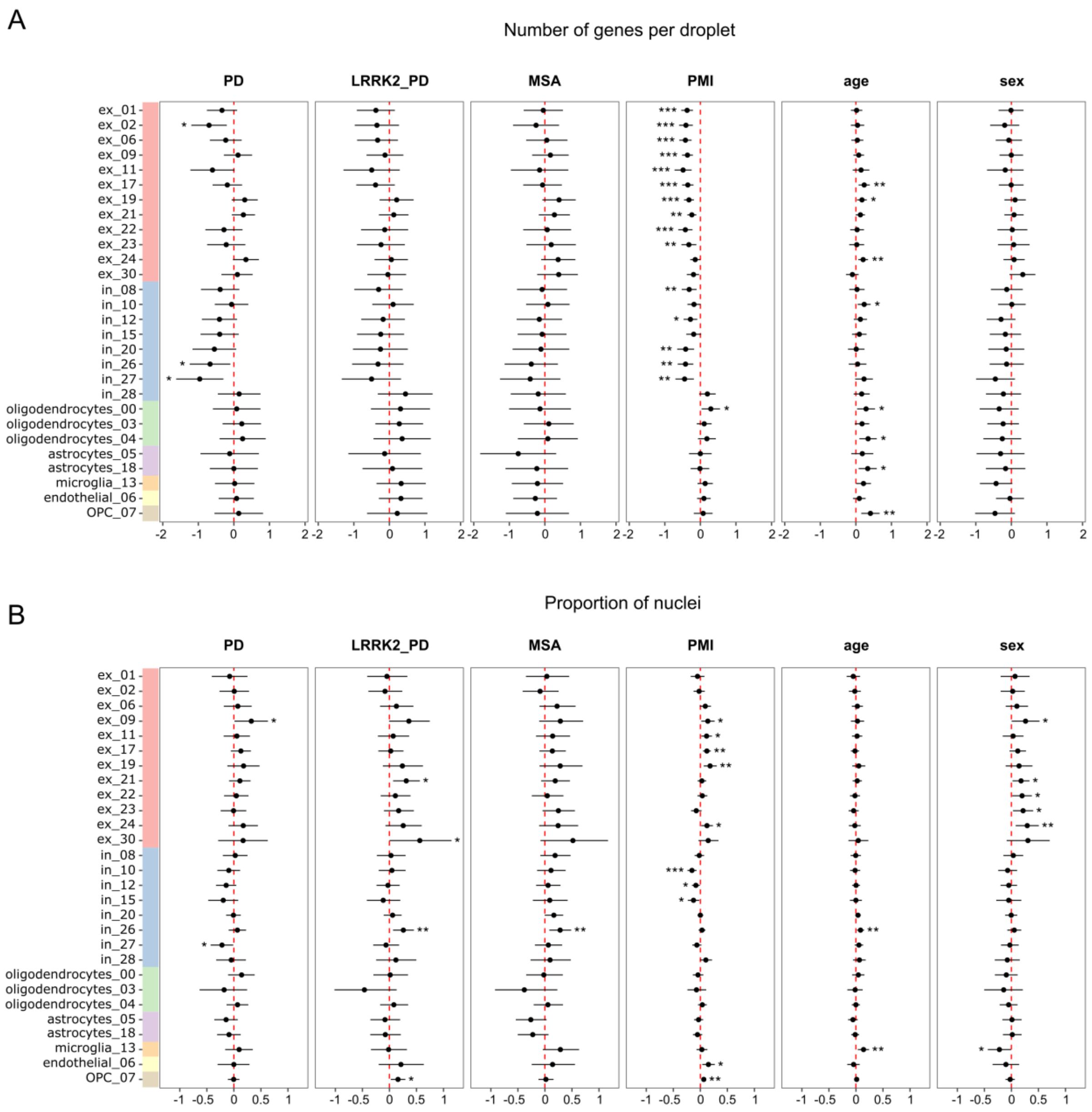


**Supplementary Figure 5**

**Cell type-specific association of demographic variables with the number of expressed genes and the number of captured nuclei.** Coefficients for the effects of each demographic variable (disease status, PMI, age and sex) on the number of expressed genes (A) and the proportion of nuclei (B) in each cell type cluster identified in the snRNA-seq. **A:** Scaled log fold-change in the number of features (genes) per droplet. Coefficients were calculated with a linear mixed model with disease status, PMI, age and sex as fixed effects and individual as a random effect. **B:** Scaled log fold-change in the proportion of nuclei. Nuclei counts were modelled as a function of disease status, PMI, age and sex with a negative binomial regression. For **A** and **B**, whiskers indicate 95% confidence interval (see Methods). “ex” – excitatory neuron; “in” – inhibitory neuron; “OPC” – oligodendrocyte precursor cell. Asterisks denote statistical significance (* $p < 0.05$; ** $p < 0.01$; *** $p < 0.001$).

### Supplementary Files

**Supplementary Table 1**

**Demographic data.** Table containing demographic variables for the samples analyzed in the study.

**Supplementary Table 2**

**Decrease in the number of expressed genes in neuronal types in iPD and LRRK2-PD.** The table shows the summary of the linear mixed model for the (log) number of genes per droplet as a function of demographic variables. Interaction terms are shown in bold. Estimates correspond to log fold-changes between diseases and controls for neuronal types with respect to glial cells. In addition to the shown fixed effects, individual and PMI were modelled as random effects (random intercept for individual and random intercept and slope for PMI).

**Supplementary Data 1**

**MAST differential gene expression output for iPD *vs* Controls.** Results from the differential gene expression analyses carried out using the MAST framework for the snRNA-seq dataset comparing iPD vs Controls. Each sheet corresponds to a cell type cluster. Statistically significant results (FDR < 0.05) are highlighted in yellow.

**Supplementary Data 2**

**MAST differential gene expression output for LRRK2-PD *vs* Controls.** Results from the differential gene expression analyses carried out using the MAST framework for the snRNA-seq dataset comparing LRRK2-PD *vs* Controls. Each sheet corresponds to a cell type cluster. Statistically significant results (FDR < 0.05) are highlighted in yellow.

**Supplementary Data 3**

**MAST differential gene expression output for MSA *vs* Controls.** Results from the differential gene expression analyses carried out using the MAST framework for the snRNA-seq dataset comparing MSA *vs* Controls. Each sheet corresponds to a cell type cluster. Statistically significant results (FDR < 0.05) are highlighted in yellow.

**Supplementary Data 4**

**Geneset enrichment analyses for the disease-specific differentially expressed genes.** Results from the geneset enrichment analysis for the differentially expressed genes corresponding to the three diseases (iPD, LRRK2-PD and MSA) compared to Controls. Sheets correspond to different Gene Ontology categories and contrasts. GO_BP: Biological Process; GO_CC: Cellular Compartment; GO_MF: Molecular Function. In each table, the “ct” column contains the cell type cluster membership.

**Supplementary Data 5**

**Geneset enrichment analysis for the GWAS candidates.** Results from the geneset enrichment analysis using the .gene list of candidates from Nalls *et al*[2]. for all the disease-specific differentially expressed genes. The "ct" column contains the cell type cluster membership, the "diagnosis" column contains the disease (iPD, LRRK2-PD, MSA).

**Supplementary Data 6**

**MAST differential gene expression output for Lewy pathology in LRRK2-PD patients.** Results from the differential gene expression analyses carried out using the MAST framework for the snRNA-seq dataset comparing LRRK2-PD samples with Lewy pathology (Braak 4-5) *vs* LRRK2-PD samples without Lewy pathology (Braak stage 0). Each sheet corresponds to a cell type cluster. Statistically significant results (FDR < 0.05) are highlighted in yellow.

**Supplementary Data 7**

**Geneset enrichment analyses for the Lewy pathology in LRRK2-PD patients.** Results from the geneset enrichment analysis for the differentially expressed genes corresponding to the comparison between LRRK2-PD samples with Lewy pathology (Braak 4-5) *vs* LRRK2-PD samples without Lewy pathology (Braak stage 0). Sheets correspond to different Gene Ontology categories. GO_BP: Biological Process; GO_CC: Cellular Compartment; GO_MF: Molecular Function. In each table, the "ct" column contains the cell type cluster membership.

**Supplementary Data 8**

**Geneset enrichment analyses for the overlapping genes between Lewy pathology in LRRK2-PD patients and disease-specific genes in iPD.** Results from the geneset enrichment analysis for the overlapping genes between the Lewy pathology contrast within LRRK2-PD patients and the iPD-specific differentially expressed genes. Sheets correspond to different Gene Ontology categories. GO_BP: Biological Process; GO_CC: Cellular Compartment; GO_MF: Molecular Function. In each table, the "ct" column contains the cell type cluster membership.

**Supplementary Data 9**

**MAST differential gene expression output for Lewy pathology in iPD patients.** Results from the differential gene expression analyses carried out using the MAST framework for the snRNA-seq dataset comparing iPD samples with varying levels of Lewy pathology (Braak 4-6). Each sheet corresponds to a cell type cluster. Statistically significant results (FDR < 0.05) are highlighted in yellow.
